# Translation of a human induced pluripotent stem cell-derived ovarian support cell product to a Phase 3 enabling clinical grade product for *in vitro* fertilization treatment

**DOI:** 10.1101/2025.04.02.25324702

**Authors:** Bruna Paulsen, Ferran Barrachina, Sabrina Piechota, Alexander D. Noblett, Mark Johnson, Simone Kats, Cassandra Lew, Maria Marchante, Alexandra B. Figueroa, Itzel Garcia Granada, Elizabeth Ingalls Lopez, Erick Martinez Martinez, Paula Ricra, Camila Carlos, Jazmin Meza, Wendy Montanchez, Pilar Pino, Cesar Reategui, Enrique Noriega, Alicia Elias, Luis Noriega-Portella, Gus Haddad, Dina Radenkovic, Eugenia Moran, Pamela Villanueva, Jose Guiterrez, Luis Guzman, Pietro Bortolleto, David F. Albertini, Michel De Vos, Christian C. Kramme

## Abstract

Human induced pluripotent stem cells (hiPSCs) show great promise in the development of novel strategies to mitigate reproductive diseases and promote successful reproductive outcomes. Recently, a novel approach for the fast and efficient differentiation of ovarian support cells (OSCs) to generate a versatile platform for basic research and clinical applications was demonstrated. This study details the clinical process development and application of an OSC product, known as *Fertilo*, to improve the *in vitro* maturation (IVM) of human oocytes, a method referred to as OSC-IVM. First, transcription factor (TF) mediated OSC differentiation using research-grade raw materials was shown to produce granulosa-like cells that improve the MII maturation rate of human oocytes. To support clinical application, several raw material upgrades were initiated, including substitution of the differentiation matrix with a higher-quality alternative, laminin-521, and the generation of a clinically suitable hiPSC seed bank and master cell bank. Single cell RNA sequencing of OSCs generated using the updated protocol for clinical translation demonstrated the consistency and reproducibility of cellular outcomes. Next, analytical release testing of the clinical product was performed and a murine oocyte maturation assay was developed to establish the potency of OSCs for use in OSC-IVM. Finally, the qualified *Fertilo* product was applied in a two-phase longitudinal cohort analysis, with the results showing improvement in key outcomes compared to traditional IVM treatment. Our findings demonstrate the first-time clinical development and application of an hiPSC-derived product to improve reproductive outcomes after IVM and advance women’s health.

## Introduction

Reproductive diseases have a significant impact on human health, particularly affecting women’s well-being. Conditions like endometriosis, polycystic ovary syndrome, and infertility are substantial public health concerns, each affecting over 10% of the population and influencing women’s health and their lifestyles (*1–3*). These conditions, which are often intertwined with comorbidities, lead to increased healthcare utilization and economic burden (*4*). Despite the high prevalence and clinical significance of these reproductive diseases, recent decades have been marked by a lack of funding and therapeutic development (*5, 6*), leading to healthcare disparities and hindering innovation.

Emerging evidence showcasing the use of human induced pluripotent stem cells (hiPSCs) in generating models for a wide range of applications provides prospects for the management of reproductive health conditions. hiPSCs have the potential to be differentiated into any cell type in the body, including ovarian support cells (OSCs) bearing phenotypic and functional similarities to somatic granulosa cells within the ovarian follicle (*7–10*). A recent study demonstrated that by modulating the expression of key transcription factors (TFs) involved in granulosa cell specification, the differentiation of hiPSCs could be orchestrated to lead to the efficient generation of mature ovarian cell types (*8*). Differentiated OSCs mimic the ovarian environment *in vitro* and therefore, constitute a platform with a plethora of applications from investigation of the basic understanding of gamete-somatic cell interactions to applications of clinical interest, such as the *in vitro* maturation (IVM) of oocytes, *in vitro* gametogenesis, disease modeling, and drug screening. A recent study demonstrated the potency of hiPSC-derived OSCs in enhancing oocyte maturation and euploid blastocyst formation via a method called OSC-IVM (*11, 12*).

IVM is a mild-approach assisted reproductive technology (ART) and has the potential to allow patients to undergo oocyte cryopreservation and *in vitro* fertilization (IVF) through a safer procedure due to the significantly lower requirement of gonadotropins for ovarian hyperstimulation. However, despite being less expensive and painful compared to standard of care procedures (*13*), IVM has been historically limited by poor outcomes, which has motivated the search for solutions to overcome these challenges to increase access to fertility treatments (*14–16*). The ability to recreate the ovarian microenvironment in a dish, by direct co-culture of OSCs and oocytes, offers an opportunity to physiologically mimic the dynamic environment necessary to support oocyte maturation *in vitro* and improve the outcomes associated with IVM (*11, 12, 17*). Nevertheless, clinical application of allogeneic OSCs for *ex vivo* gamete co-culture requires a meticulous approach to address several process challenges, namely utilization of a clinically appropriate hiPSC line and the highest quality available raw materials, reproducibility in differentiation outcomes, and consistency in potency in terms of promoting human oocyte maturation.

This manuscript details the strategic development of an hiPSC-derived product, known as *Fertilo*, through clinical translation. Following initial proof of concept experiments, product development studies progressed with the implementation of higher grade raw materials in the manufacturing of OSCs and the characterization of resultant OSC populations. After validating that the differentiation process led to an efficacious and consistent population of OSCs, several lots of OSCs were generated and demonstrated to be suitable for clinical use based on analytical release testing and a murine oocyte maturation assay to assess potency. Finally, OSCs were applied clinically via a first-in-human evaluation of OSC-IVM, leading to improved rates of euploid blastocyst formation and ongoing pregnancy compared to standard of care IVM, in addition to the first successful, healthy live birth in humans. This study is the first to demonstrate the end-to-end process of the manufacturing, product release, and application of a product developed from hiPSCs that has progressed to registrational Phase 3 clinical trials in the United States. Notably, this product is also the first hiPSC-derived therapy developed for use in *in vitro* fertilization (IVF).

## Results

### Transcription factor-mediated hiPSC differentiation generates OSCs that improve the rate of MII oocyte maturation

*Fertilo* is a cell based product composed of engineered OSCs, derived from hiPSCs, which are applied in the IVM of human oocytes in a method called OSC-IVM. To guide clinical translation of the *Fertilo* product and establish good manufacturing practice (GMP) and preclinical safety and efficacy assessment using a Quality by Design (QbD) approach, a Quality Target Product Profile (QTPP) was generated (**Table 1**). To refine and improve the QTPP, a series of process development steps and preclinical studies were performed to establish the differentiation protocol, efficacy in a therapeutic model, and the toxicological profile (**Table 2**).

**Table 1:**
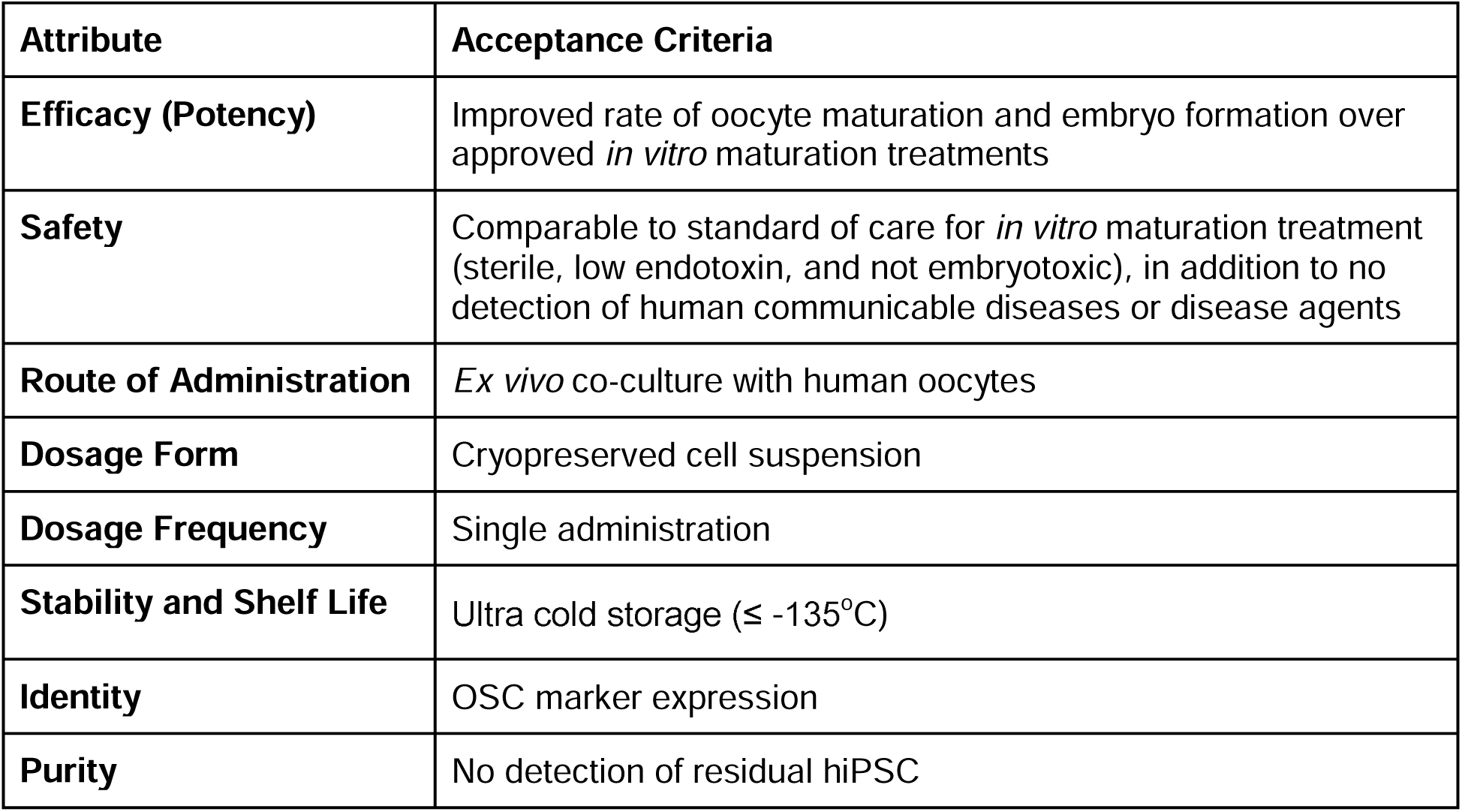
Quality Target Product Profile of *Fertilo*.

**Table 2:**
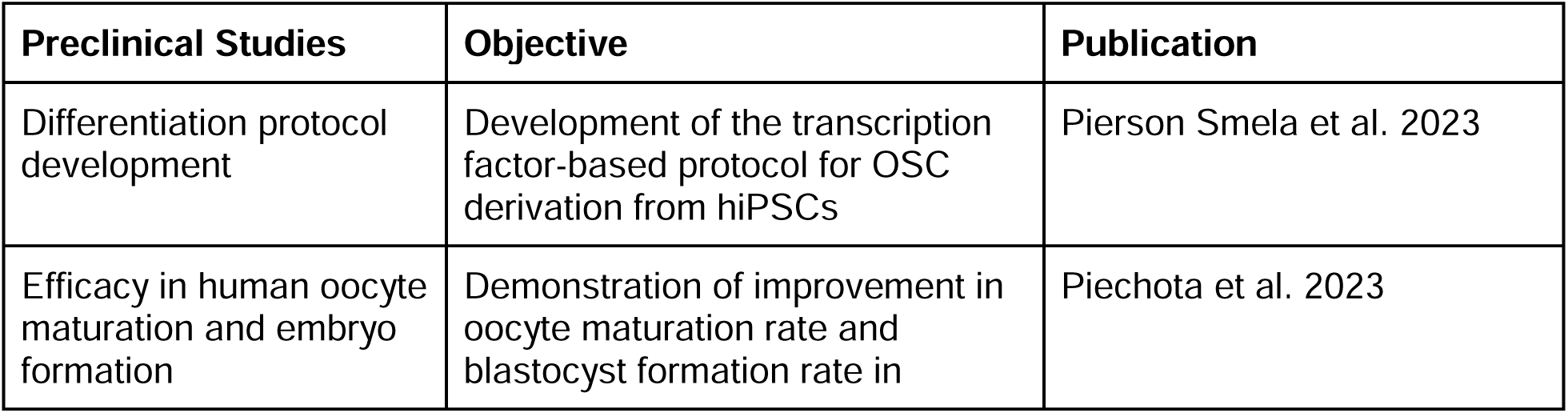
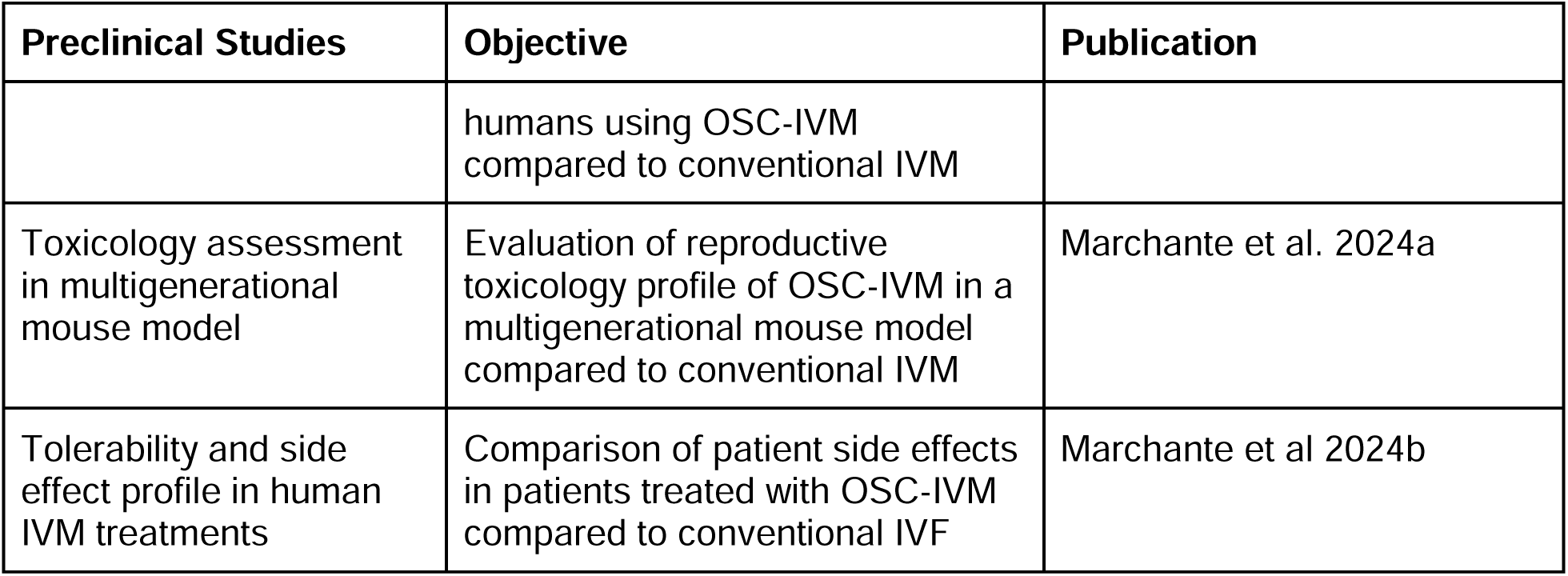
Preclinical Studies of OSC-IVM.

Previous studies have demonstrated that modulating the expression of key transcription factors involved in granulosa cell specification can drive the differentiation of hiPSCs toward mature ovarian cell types, referred to as OSCs, that carry phenotypic and functional similarities to somatic granulosa cells of the endogenous ovary (*8*). To evaluate the phenotypic consistency of the previously described research use only (RUO)-hiPSC line harboring three inducible transcription factors (*NR5A1*, *RUNX2*, and *GATA4*), which was used to generate functional OSCs in previous studies (*8*), six independent batches of OSCs produced over eight months by multiple operators were compared (**Table S1**). After five days of induction on matrigel (M), RUO-hiPSCs multiplied 5.63±2.85 times and acquired morphological features that resembled human granulosa cells, such as clusters of cells with spiky edges and granules in the cell body (**Fig. 1A**). Differentiated RUO-OSCs also expressed CD82, a marker associated with granulosa cell fate (*8, 18*), indicating successful differentiation into the desired cell type (**Fig. 1B**).

**Fig. 1:**
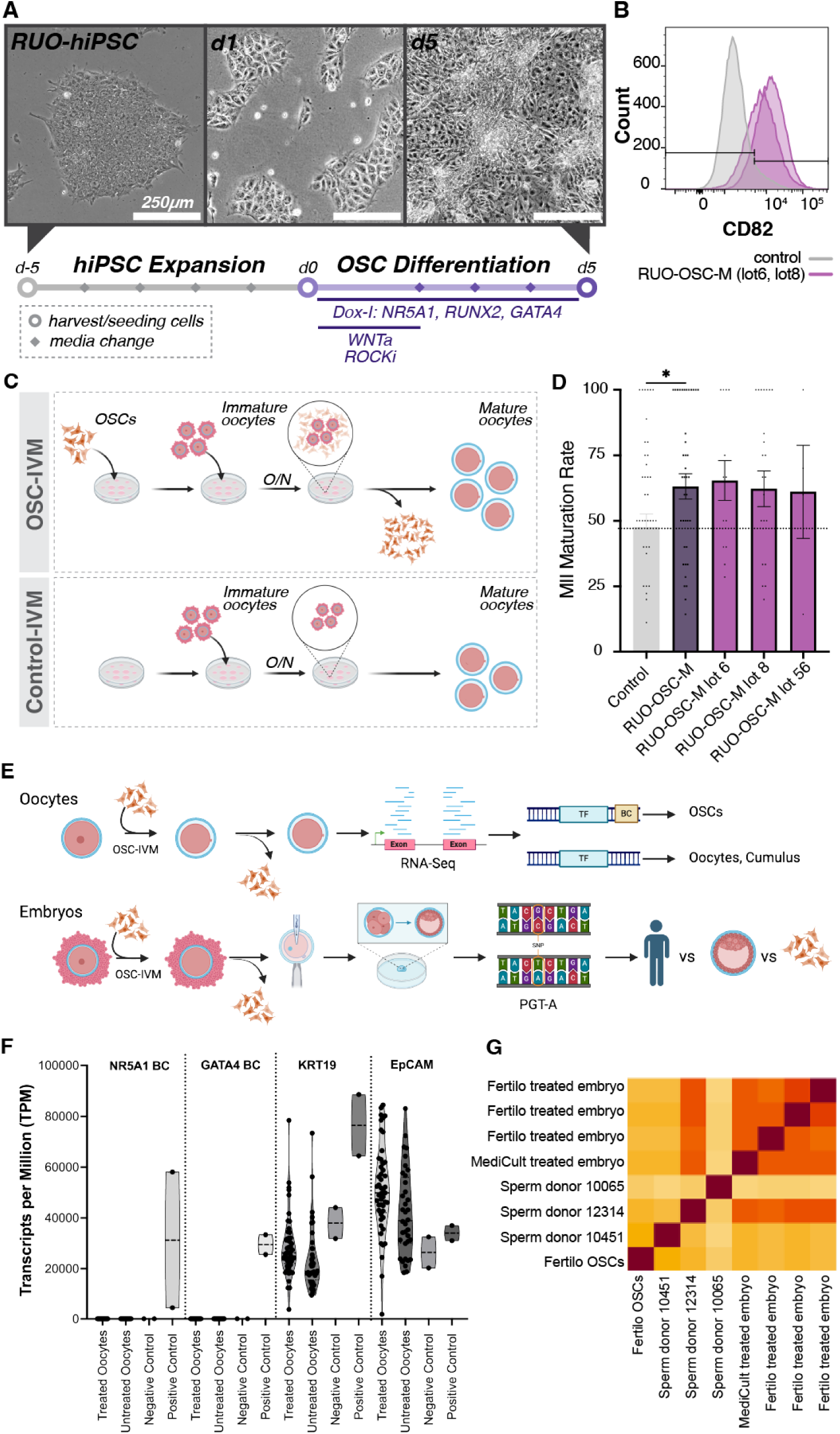
Transcription factor mediated hiPSC differentiation to generate OSCs that improve the rate of MII oocyte maturation. A) Timeline and representative images of RUO-hiPSC expansion and OSC differentiation onto matrigel (M). Images on day 5 of hiPSC expansion and days 1 and 5 of OSC differentiation. Scale bar, 250 μm. B) Flow cytometry analysis of the granulosa cell marker, CD82, in RUO-OSC-M lots and the negative (unstained) control. C) Schematic representation of the two *in vitro* maturation (IVM) conditions: the Control-IVM group containing IVM media only versus the OSC-IVM group constituted by IVM media supplemented with OSCs. D) Quantification of MII Maturation Rate in Control-IVM (grey) vs OSC-IVM groups. “RUO-OSC-M” displays the combined oocyte maturation rates of 3 separate OSC batches (RUO-OSC-M lots 6, 8, and 56). Data are shown as the mean ± SEM (*p*=0.029, lot 6 vs control:1.37, lot 8 vs control: 1.31, lot 56 vs control: 1.28). E) Flowchart demonstrating the testing strategy to verify OSC removal from post-treatment human oocytes and embryos. Barcode-seq analysis was performed on post-treatment oocytes and SNP-based PGT-A analysis was performed on post-treatment embryos. F) Barcode-seq analysis of 48 post-treatment oocytes. Analysis of the presence of barcodes (as transcripts per million) for OSCs with synthetic TFs and the reference genes KRT19 and EpCAM. Analysis was performed using both OSC line data (bulk RNA-seq) and post-IVM oocyte cell data (low input RNA-seq). Post-IVM oocytes show no detectable transcript count of either synthetic TF barcode region, while the OSC line controls show the clear presence of active barcode transcripts while reference cumulus cell controls show no detection. Transcript capture of reference genes show high read depth and quality library preparation. G) SNP-based PGT-A analysis of post-treatment embryos, sperm donor samples, and *Fertilo* OSCs. Patient oocytes were treated with *Fertilo* or IVM Media. Mature oocytes were fertilized using sperm from Sperm Donor 12314, resulting in 4 blastocysts sent for PGT-A analysis, 3 derived from *Fertilo* treatment and 1 derived from IVM treatment. SNP detection via PGT-A analysis was performed and included analysis of negative controls (Sperm Donor 10451 and Sperm Donor 10065), as well as a sample of *Fertilo* OSCs (GTO-101-Lot-006) and the fertilization partner (Sperm Donor 12314). No evidence of residual *Fertilo* OSCs, measured via unique SNP detection, was found in any *Fertilo* treated embryos. The assay correctly identified the fertilization partner, with non-partner sperm showing no evidence of overlap with embryo samples. There was no difference in SNP profiles between the IVM derived embryo and the *Fertilo* derived embryos.

The RUO-OSCs were then applied for IVM co-culture using human oocytes as a measure of product potency and safety. Following IVM, MII oocyte maturation rate was assessed, and all tested RUO-OSC-M batches successfully led to higher MII oocyte maturation rates compared to the control group (*p*=0.029, lot 6/control:1.37, lot 8/control: 1.31, lot 56/control: 1.28) (**Fig. 1C,D**). A key safety aspect of *Fertilo* use is the exclusive *ex vivo* nature of application and the elimination of OSCs, which prevents allogeneic cell transfer to patients or progeny. To verify OSC removal, we analyzed human oocytes and embryos post-treatment using next-generation sequencing (NGS) to identify any residual OSCs (**Fig. 1E**) (*12*). hiPSC-derived OSCs contain a unique 20-basepair barcode in the 3’ UTR of exogenous TFs that enables barcode capture using RNA-sequencing to distinguish the residual presence of OSCs versus cumulus cells in oocytes (*19*). Barcode-seq analysis of 48 post-treatment oocytes showed no residual OSCs, confirmed by the high specificity and sensitivity in controls (positive: OSCs, n=2; negative: untreated oocytes, n=36; cumulus cells, n=2) (**Fig. 1F**). Additionally, single nucleotide polymorphism (SNP)-based preimplantation genetic testing for aneuploidy (PGT-A) of biopsied human blastocysts can distinguish between maternal and paternal genetic material versus OSC genetic material in the embryo. PGT-A analysis of 26 human blastocysts derived from OSC-treated oocytes and 13 control untreated embryos, compared to parental and OSCs genomes, revealed no OSC-derived genetic material in the resultant embryos (**Fig. 1G**). These findings support the safety of OSC-IVM use, demonstrating the minimal risk of OSC transfer when used appropriately.

### Translation of the protocol towards clinical manufacturing with high-quality clinically suitable raw materials

Despite the generation of OSCs with the desired identity (CD82 expression) and functional profiles (improvement in MII maturation rate of human oocytes), in addition to verification that no residual OSCs were detected beyond application, the raw materials utilized throughout the process were not of the highest quality available and therefore not suitable for manufacturing a clinical product. Hence, a thorough evaluation was necessary to determine whether substituting raw materials with higher quality alternatives would alter phenotypic attributes and potency.

To ensure a seamless transition to more suitable raw materials without compromising the key quality attributes of the final product, particularly purity and potency, a risk assessment was conducted alongside a systematic evaluation of differentiation outcomes. A list of critical factors, including basal media, serum replacements, small molecules, and growth factors/morphogens, was compiled from the literature, and appropriate test concentrations were identified (**Table S2**). To investigate the effects of multiple variables at once, a Design of Experiments (DOE) approach was applied to optimize FOXL2 expression and viability; FOXL2 indicates OSC specification and viability screens for environmental factors that are otherwise unworkable in manufacturing (**Fig. 2A**). The results demonstrated that substrate had a clear and strong influence on FOXL2 expression (**Fig. 2B**, *p* ≤ 0.01), far greater than any other factors.

**Fig. 2:**
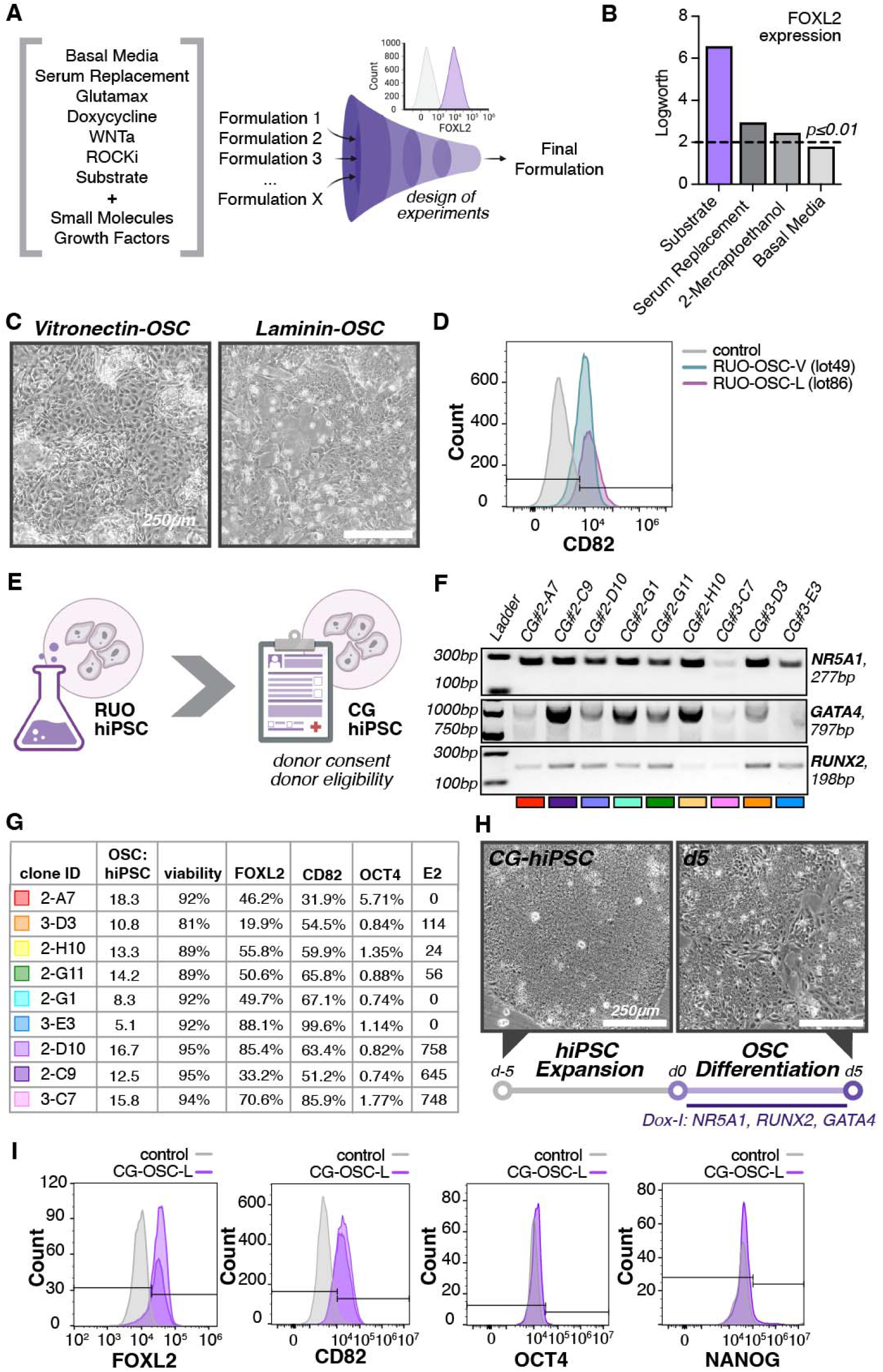
Translation of the protocol towards clinical manufacturing using high-quality clinically suitable raw materials. A) Schematic of the design of experiments (DOE) strategy to optimize the manufacturing process. B) Barplot of logworth values of DOE main effect on FOXL2 expression. The dashed line indicates p≤0.01. C) Images of OSCs in culture on day 5 of differentiation carried out on vitronectin vs laminin-521. Scale bar, 250 μm. D) Flow cytometry analysis of the expression of CD82 in the control, RUO-OSC-V and RUO-OSC-L. E) Schematics of requirements for generation of a clinical-grade (CG)-hiPSC line. F) Image of genotyping PCR gel confirming the presence of all three transcription factors (*NR5A1*, *GATA4*, and *RUNX2*) in each of the 9 clones. G) Chart displaying the 9 clones and 4 markers: FOXL2, CD82, OCT4, and E2. The chart displays the ratio of the presence of OSC markers (FOXL2 and CD82) in relation to the hiPSC markers (OCT4 and E2) in each clone, the viability of each sample, and the amount of each marker present in each sample as a percentage. H) Images of clinical grade OSCs (lot 90) grown on laminin on day 5 of hiPSC expansion and day 5 of OSC differentiation. Scale bar, 250 μm. I) Flow cytometry analysis of OSC markers: FOXL2, and CD82. Expression levels of these markers were evaluated in CG-OSC-L against the negative control.

For this reason, a detailed analysis of alternative substrates was a key focus in this stage of process development. Indeed, one of the original raw materials of highest complexity was Matrigel, a cell substrate derived from Engelbreth-Holm-Swarm mouse sarcoma cells that contains multiple extracellular matrix components of tissue basement membranes. Due to its nature of production, Matrigel is also subject to significant lot-to-lot variability, which can impact the overall reproducibility and purity of the final product. The substrate alternatives included commercially available, animal origin-free extracellular matrix substrates appropriate for the translation of clinical cell therapies—laminin and vitronectin. Two different physiologically relevant laminin isoforms, laminin-521 and laminin-511, were evaluated by seeding hiPSCs on each matrix, inducing differentiation, and evaluating cells for FOXL2 expression and metrics of workability. The results indicated that laminin-521 best met the targeted outcomes (**Fig. S1**).

To evaluate the substrates, differentiation of the RUO-hiPSC line was performed using the same method aside from utilization of the alternative substrates. Initial assessment of cellular morphology during the differentiation process indicated subtle differences between both groups (**Fig. 2C**). Vitronectin-OSCs (RUO-OSC-V) exhibited a larger cell body and organized themselves in sparse clusters of cells, while laminin-OSCs (RUO-OSC-L) were smaller and organized in compact groups (**Fig. 2C**). Expression of the granulosa-cell marker CD82 was present in both OSC groups (**Fig. 2D**). Analytical testing of the different OSC lots demonstrated that RUO-OSC-L lots had an increased percentage of cells expressing the desired OSC markers, increased cell viability, and significantly reduced expression of the hiPSC marker, TRA-1-60, compared to RUO-OSC-V lots, indicating a more efficient and pure differentiation process with laminin-521 (**Table 3**).

**Table 3:** Analytical Test Results for RUO-OSC-V and RUO-OSC-L lots.

| Cell Line | OSC Marker (CD82+) | Viability | Residual hiPSCs (TRA-1-60+) |
| --- | --- | --- | --- |
| RUO-OSC-V lot 41 | 51.1% | 81.7% | 31.1% |
| RUO-OSC-V lot 49 | 57.7% | 82.8% | 72.5% |
| RUO-OSC-L lot 77 | 50.8% | 86.5% | 3.5% |
| RUO-OSC-L lot 86 | 87.4% | 86.2% | 1.3% |

To generate a clinical-grade (CG) starting material, an hiPSC line derived from an allogeneic female donor with proper consent and eligibility was obtained from a commercial vendor (Reprocell USA). This CG-hiPSC line, whose donor eligibility was in accordance with 21 CFR 1271 Subpart C, was genetically engineered using the same strategy developed for the RUO-hiPSC line (**Fig. 2E**). In short, the donor hiPSC line was engineered to harbor inducible versions of three TFs, *NR5A1*, *RUNX2*, and *GATA4*; when these TFs are expressed, the resultant cells have a transcriptomic profile similar to granulosa cells, produce estradiol (E2) in response to follicle stimulating hormone (FSH), and support the maturation of germ cells (*8*). Individual clones were generated by limiting the dilution of the pooled engineered population and expanding into seed banks. All expanded clones were subjected to initial screening by genotyping PCR to confirm integration of the three TFs, and nine were found to harbor the desired TF combination (**Fig. 2F**). The performance of these clones was then assessed via flow cytometry based on their ability to differentiate into OSCs, as indicated by FOXL2 and CD82 expression, with few residual hiPSCs, as indicated by OCT4 expression, after 5 days of differentiation. All clones were positive for both OSC markers with low or null hiPSC marker expression, indicating successful differentiation (**Fig. 2G, Fig. S2A**).

On the fifth day after differentiation, the clones were evaluated for steroidogenic potential by measuring estradiol (E2) production in response to follicle-stimulating hormone (FSH, #2), androstenedione (A4, #3), or a combination of both (FSH+A4, #4) for 24 hours (**Fig. S2B**). The clones responded differently to (FSH+A4), with clones 2-D10 and 3-C7 showing the greatest steroidogenic responses (**Fig. 2G**). In addition, bulk RNA-seq of clones was performed, and all clones robustly expressed most well-known granulosa cell markers (*FOXL2*, *STAR*, and *GJA5*), as well as genes related to important signaling pathways associated with oocyte-granulosa-cell interactions (*NOTCH3*, *HES1*, *ID3*, and *KITLG*) (**Fig. S2C**) (*15*). Overall, clone 2-D10 was the top candidate (hereafter referred to as CG-hiPSC) based on positive FOXL2 and CD82 expression, reduced levels of residual hiPSCs, high E2 production, high cell viability at harvest, and the highest ratio of OSCs to hiPSCs seeded at the start of differentiation (OSC:hiPSC) (**Fig. 2G**).

In addition to updating the substrate and starting material, the process was further optimized by replacing raw materials for higher-quality alternatives and the use of a T-flask instead of an open well plate system. Then, three independent CG-OSC batches (CG-OSC-L lot 88, lot 90, and lot 116) were generated leveraging the updated protocol. To evaluate the potential for scalability, while the first two lots of CG-OSCs (lot 88 and lot 90) were differentiated onto 55 cm^2^ dishes (**Table S1**), CG-OSC-L lot 116 was differentiated onto 175 cm^2^ dishes. In all lots, cell morphology upon differentiation was characterized by small cells with granules in the cell body, and cells were tightly packed into clusters with spiky edges (**Fig. 2H**). The identity of OSCs was confirmed based on expression of the markers FOXL2 and CD82, and purity was verified based on the absence of hiPSC marker expression (**Fig. 2I, Fig. S3**). Viability at harvest also remained high, averaging 97.17±0.56% among the three batches of CG-OSCs. The ratio of OSC:hiPSC was similar to the ratio achieved with the RUO-hiPSC line when differentiated over laminin-521 (14.83±4.48), averaging 11.41±2.19 for the first two batches (lot 88 and lot 90, **Table S1**). Interestingly, differentiation of CG-OSC-L lot 116 onto larger dishes (175 cm^2^) led to a considerably higher OSC:hiPSC ratio (30.7, **Table S1**), suggesting notable scalability potential.

### Transcription factor mediated hiPSC differentiation using clinical grade materials leads to OSCs with consistent cellular outcomes

To assess the reproducibility of cellular outcomes, as well as consistency throughout process development, scRNA-seq was performed on six batches of RUO-OSC-M, three batches of RUO-OSC-V, two batches of RUO-OSC-L, and two batches of CG-OSC-L (**Fig. 3A**). The scRNA-seq analysis revealed three distinct profile clusters that were classified as less mature granulosa-like cells (Early GC), more mature granulosa-like cells (GC), and “Others”. The Early GC and GC clusters robustly expressed markers of granulosa cell fate (**Fig. S4A**), with Early GCs also exhibiting expression of pre-defined preGC-I/II marker genes (**Fig. S4B**) (*20*). Those classified as “Others” were enriched in mitochondrial and ribosomal gene expression, typically associated with poor cell quality and a common artifact of scRNA-seq experiments (*21*). These datasets were also evaluated using gene signature scores established in a published transcriptome landscape of human folliculogenesis (*18*). The gene clusters identified in this study had clear signature scores for ‘Antral GC’ compared to ‘Pre-ovulatory GC’ (**Fig. S4C**), with multiple signature genes present in different clusters.

**Fig. 3:**
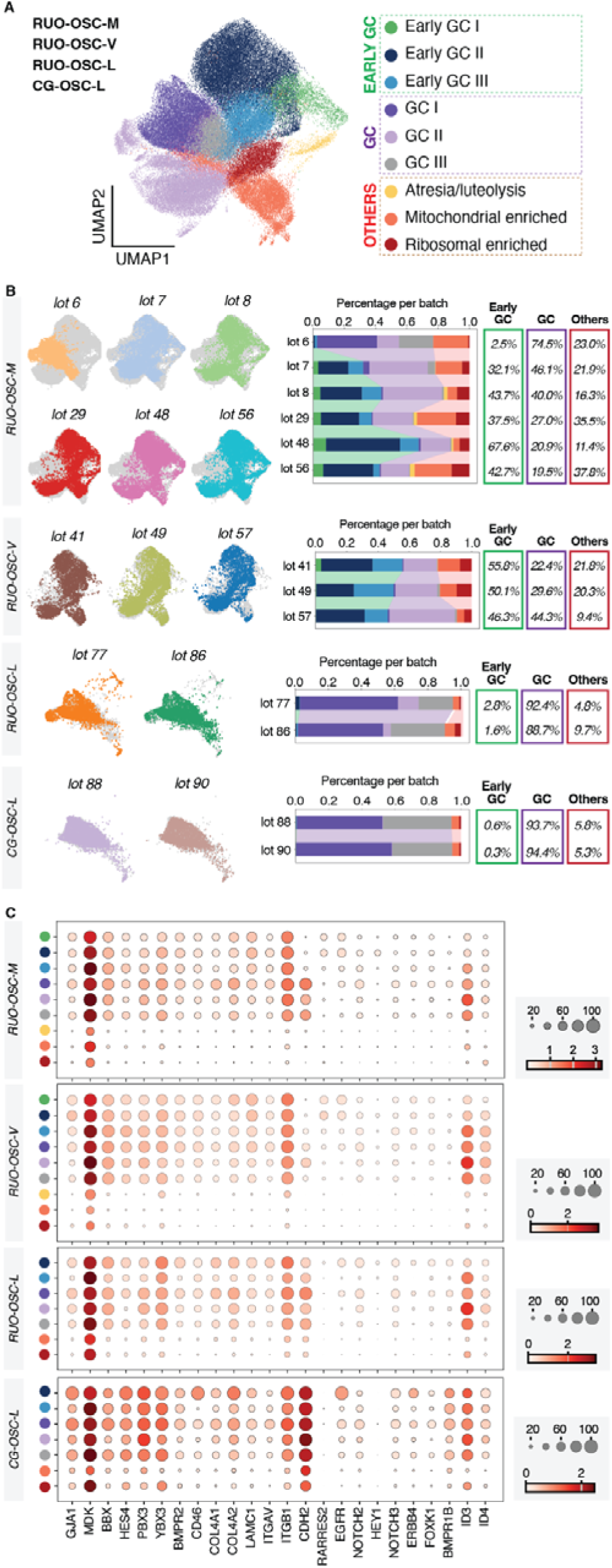
Transcription factor mediated hiPSC differentiation using clinical grade materials leads to OSCs with consistent cellular outcomes. A) UMAP projection depicting all OSC subsets, including the RUO-OSC-M, RUO-OSC-V, RUO-OSC-L, and CG-OSC-L subsets. B) UMAP projections depicting each individual lot within the RUO-OSC-M, RUO-OSC-V, RUO-OSC-L, and CG-OSC-L subsets. Next to projections for each lot, a stacked bar plot depicts the amount of each cluster type found in each lot relative to each respective subset. The colors correspond to the UMAP cluster colors found in Fig. 3A. Overall percentages per group are given to the right of the barplot. C) Dotplot depicting the expression of granulosa cell marker genes consistently expressed across GC clusters (GC consistent) and granulosa cell marker genes defined in previous studies (GC) in the RUO-OSC-M, RUO-OSC-V, RUO-OSC-L, and CG-OSC-L subsets (*19*). The RUO-OSC-M dotplot Scale represents ‘Mean expression in groups’ and the circles represent ‘Fraction of cells in group (%)’.

The scRNA-seq data per individual lot showed that the RUO-OSC-M lots generated cells with variable distribution among the three major classes, while the RUO-OSC-V and RUO-OSC-L lots generated cells with more consistent outcomes, exhibiting an increased percentage of Early GCs and GCs, respectively (**Fig. 3B, Fig. S5**). Finally, the CG-OSC-L lots had a consistent high proportion of the desired mature population of GCs (>93%), with a decrease from 22% to <6% of low quality cells (‘Others’) compared to the initial batches (**Fig. 3B, Fig. S5**). These data were also analyzed to evaluate the expression of GC markers in the different batches of OSCs (RUO-OSC-M, RUO-OSC-V, RUO-OSC-L, and RUO-OSC-M) (*20*). The results demonstrated that as the process was developed with higher grade raw materials, there was an increase in the expression levels of GC markers, indicating a purer and more efficient differentiation process (**Fig. 3C**). Furthermore, the functionality of the resultant OSCs in OSC-IVM was assessed, with all lots of OSCs leading to improved rates of MII maturation of human oocytes (**Fig. S6**).

To gain further insight into product development from this dataset, the scRNA-seq dataset was leveraged to investigate molecular pathways, including analysis of the gene expression of key receptor-ligand components and growth factors that could be associated with the mechanism of action of OSC-IVM. Analysis of key receptor-ligand components revealed consistent enrichment of the receptors *TGFBR1*, *BMPR2*, and *NOTCH2/3*, as well as the ligand *EFNB2*, among OSC batches, suggesting their potential involvement in OSC-IVM (**Fig. S7A**). Of note, the NOTCH signaling pathway is known to be involved in oocyte-GC crosstalk during folliculogenesis (*18*), and high levels of NOTCH2 and NOTCH3 expression in cumulus cells have been positively correlated with IVF response (*22*). Gene expression analysis of growth factors revealed the enrichment of growth factors that are well known to regulate oocyte maturation and developmental competence acquisition both *in vivo* and *in vitro*, such as *MDK*, *TGFB1*, *EGF*, *FGF2*, and *IGF2BP1/2/3*, thereby supporting a potentially paracrine mechanism of action (**Fig. S7B**) (*23–28*). Complementary proteomics analysis of OSCs after 24 hours *in vitro* compared to OSCs prior to culture (0 hour) (**Table S5**), as well as OSCs (0 hour) compared to hiPSCs (0 hour) in both genetic backgrounds (**Table S3**), was performed. The overexpressed proteins in OSCs post-24 hour culture were enriched for functional profiling terms such as “transporter activity”, “electron transfer activity”, “aerobic respiration”, and “cellular lipid metabolic process” (**Fig. S8A**); meanwhile, the proteins overexpressed in OSCs versus hiPSCs were enriched in functional profiling terms such as “cell-cell adhesion mediator activity”, “cytoskeleton organization”, and “focal adhesion” (**Fig. S8B**). Among the proteins secreted by both the RUO and CG cell lines, relevant players associated with oocyte maturation and developmental competence, such as TGFB1, EGFR, and IGF2BP1/2/3, were identified (**Fig S8C, Table S4**).

### Process robustness following optimization is demonstrated through analytical assessment of the product’s quality, safety, and purity

After confirming that the identity and purity of the final OSC product were not negatively impacted by changes in optimization during process development, two additional lots of CG-OSC-L were established to demonstrate process robustness. To ensure process consistency, analytical development was necessary to define the critical attributes required to assess potency and safety, ensuring alignment with the acceptance criteria outlined in the QTPP (**Table 1**). Since OSCs are derived from hiPSCs, their key safety considerations extend beyond the standard attributes expected for IVF-related products, such as sterility, endotoxin levels, and embryotoxicity. Additional critical attributes include residual hiPSC presence, the detection of communicable diseases, and screening for disease-related agents.

Under controlled conditions, two additional OSC lots (#180 and #182) were manufactured and compared with previously characterized CG-OSC-L lots, indicating that the cells maintained the desired characteristics, including high OSC purity and cell viability; these results demonstrated the reproducibility and consistency of the process (**Table 4**). The CG-OSC-L lots were also tested for mycoplasma, endotoxin, sterility, and human adventitious agents to ensure the safety of the clinical product.

**Table 4:** Analytical Testing of the Clinical Attributes of CG-OSC-L Lots.

| **Producti**<br>**on lot** | **OSC**<br>**Markers**<br>**(CD82+,**<br>**FOXL2+)** | **Viability** | **Residual**<br>**hiPSCs**<br>**(TRA-1-**<br>**60+)** | **Mycopla**<br>**sma** | **Endotoxin** | **Sterility** | **Adventiti**<br>**ous**<br>**Agents**<br>**(Human**<br>**Test**<br>**Panel)** |
| --- | --- | --- | --- | --- | --- | --- | --- |
| CG-<br>OSC-L<br>lot 88 | CD82+:<br>78.3%<br>FOXL2+:<br>71.2% | 90.20% | 0.01% | Neg | ≤ 0.25<br>EU/mL | NG | ND |
| CG-<br>OSC-L<br>lot 90 | CD82+:<br>74.1%<br>FOXL2+:<br>92.1% | 91.20% | 0.03% | Neg | ≤ 0.25<br>EU/mL | NG | ND |
| CG-<br>OSC-L | CD82+:<br>97.4% | 94.63% | 0.08% | Neg | ≤ 0.25 | NG | ND |

| Producti<br>on lot | OSC<br>Markers<br>(CD82+,<br>FOXL2+) | Viability | Residual<br>hiPSCs<br>(TRA-1-<br>60+) | Mycopla<br>sma | Endotoxin | Sterility | Adventiti<br>ous<br>Agents<br>(Human<br>Test<br>Panel) |
| --- | --- | --- | --- | --- | --- | --- | --- |
| lot 116 | FOXL2+:<br>99.5% |  |  |  | EU/mL |  |  |
| CG-<br>OSC-L<br>lot 180 | CD82+:<br>94.2%<br>FOXL2+:<br>99.2% | 94.37% | 0.46% | Neg | ≤ 0.25<br>EU/mL | NG | ND |
| CG-<br>OSC-L<br>lot 182 | CD82+:<br>98.5%<br>FOXL2+:<br>90.8% | 88.40% | 0.87% | Neg | ≤ 0.25<br>EU/mL | NG | ND |

Neg = negative, NG = no growth, ND = not detected

To assess the potency of CG-OSCs for *Fertilo* product release, a murine oocyte maturation assay was developed to mimic clinical application using a surrogate species (mice). In this assay, fresh immature mouse oocytes from hybrid strain B6/CBA mice at the germinal vesicle (GV) stage were collected from minimally stimulated female mice between 6 and 8 weeks of age. The immature oocytes were then subjected to IVM with IVM Media alone (Media Only Control) or in the presence of different types of cells, including OSCs. Following IVM, the matured oocytes were fertilized via intracytoplasmic sperm injection (ICSI). After 5 days, the blastocyst formation rate (BFR) was assessed as a measure of potency (**Fig. 4A**).

**Fig. 4:**
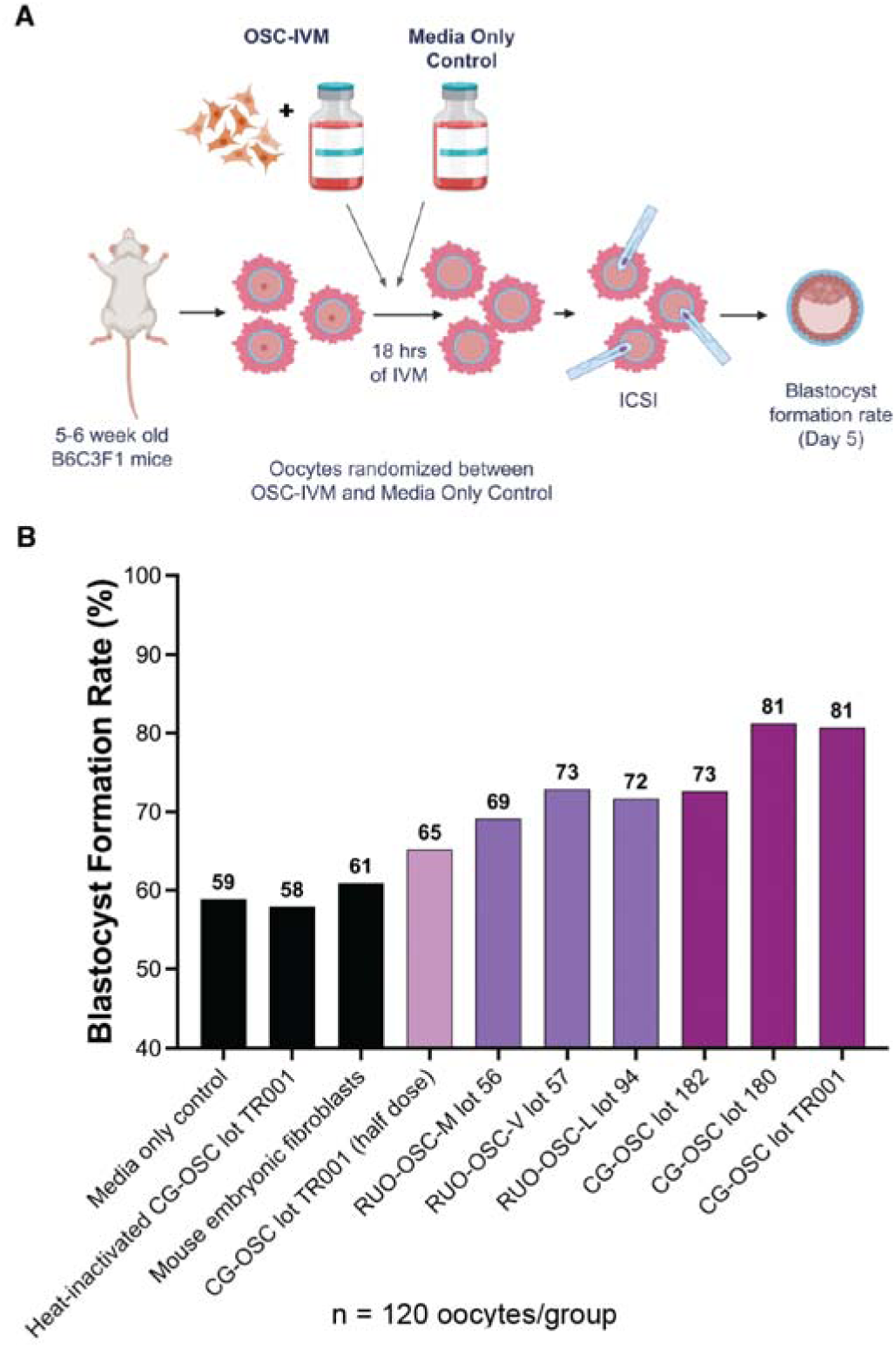
Establishment of a murine oocyte maturation assay (MOMA) for release of the clinical OSC-IVM product. A) Schematic of the MOMA study design for OSC-IVM product release. B) Blastocyst formation rate following OSC-IVM in various test conditions. The first three conditions represented by the black bars are the negative controls with no cells, inactivated cells, and alternative cells, respectively. The fourth condition is CG-OSCs with half the recommended amount of OSCs for OSC-IVM. Conditions five through seven represent OSC-IVM with RUO-OSCs, and the last three conditions represent OSC-IVM with CG-OSCs. A total of 120 oocytes were evaluated in each condition.

This study included three negative control conditions, including the media only control without cells, media with non-viable (heat-inactivated) CG-OSCs, and media with mouse embryonic fibroblasts. The BFRs were relatively consistent among these conditions (59%, 58%, and 61%, respectively), indicating that the improvement in BFR with OSC-IVM was a result of the specific activity of viable OSCs (**Fig. 4B**). Given that no differences were observed among the control conditions, the media only control (without cells) was deemed the optimal negative control; thereby eliminating the need for external cells in the control group and reducing variability. To evaluate the impact of dose on OSC-IVM efficacy, in one condition, half of the recommended number of OSCs (50,000 OSCs per 100 µL) was added during IVM co-culture. In this condition, there was approximately half of the effect on day 5 BFR compared to the full dose, demonstrating the importance of the number of OSCs in OSC-IVM (**Fig. 4B**) and the ability of the method to detect effects that are directly proportional to the concentration of OSCs (linearity). To confirm the robustness of the process, the potency of OSCs derived from different genetic backgrounds (RUO-hiPSC and CG-hiPSC) was evaluated based on BFRs following OSC-IVM. All OSC conditions (RUO-OSC-M, RUO-OSC-V, RUO-OSC-L, or CG-OSC lots) exhibited an increased BFR compared to controls, with the highest BFRs seen with the CG-OSC lots (**Fig. 4B**).

### Clinical readiness of CG-OSC-L batches for OSC-IVM application

To ensure GMP readiness, the CG-hiPSC Seed Bank, initially generated under controlled non-classified conditions, was expanded into a Master Cell Bank (MCB) under GMP-compliant conditions, following 21 CFR 210/211 regulations (**fig. S9A**). As part of this transition, aseptic process simulations (APS runs) were conducted to validate sterility, assess potential contamination risks, and confirm the robustness of the manufacturing process before full-scale GMP production (**fig. S9B**).

A comprehensive risk assessment was performed on all materials and process steps involved in the CG-hiPSC to CG-OSC manufacturing workflow. Based on the identified risks inherent to the process, targeted analytical tests were conducted to mitigate concerns and ensure process control (**Table 5**). The results demonstrated that following CG-hiPSC expansion under GMP conditions, all tests met the acceptance criteria, effectively addressing identified risks. These findings confirm that the manufacturing process had been sufficiently de-risked, supporting the progression of OSCs to GMP manufacturing and the subsequent release of GMP *Fertilo* batches for OSC-IVM application (**Table 5**).

**Table 5:**
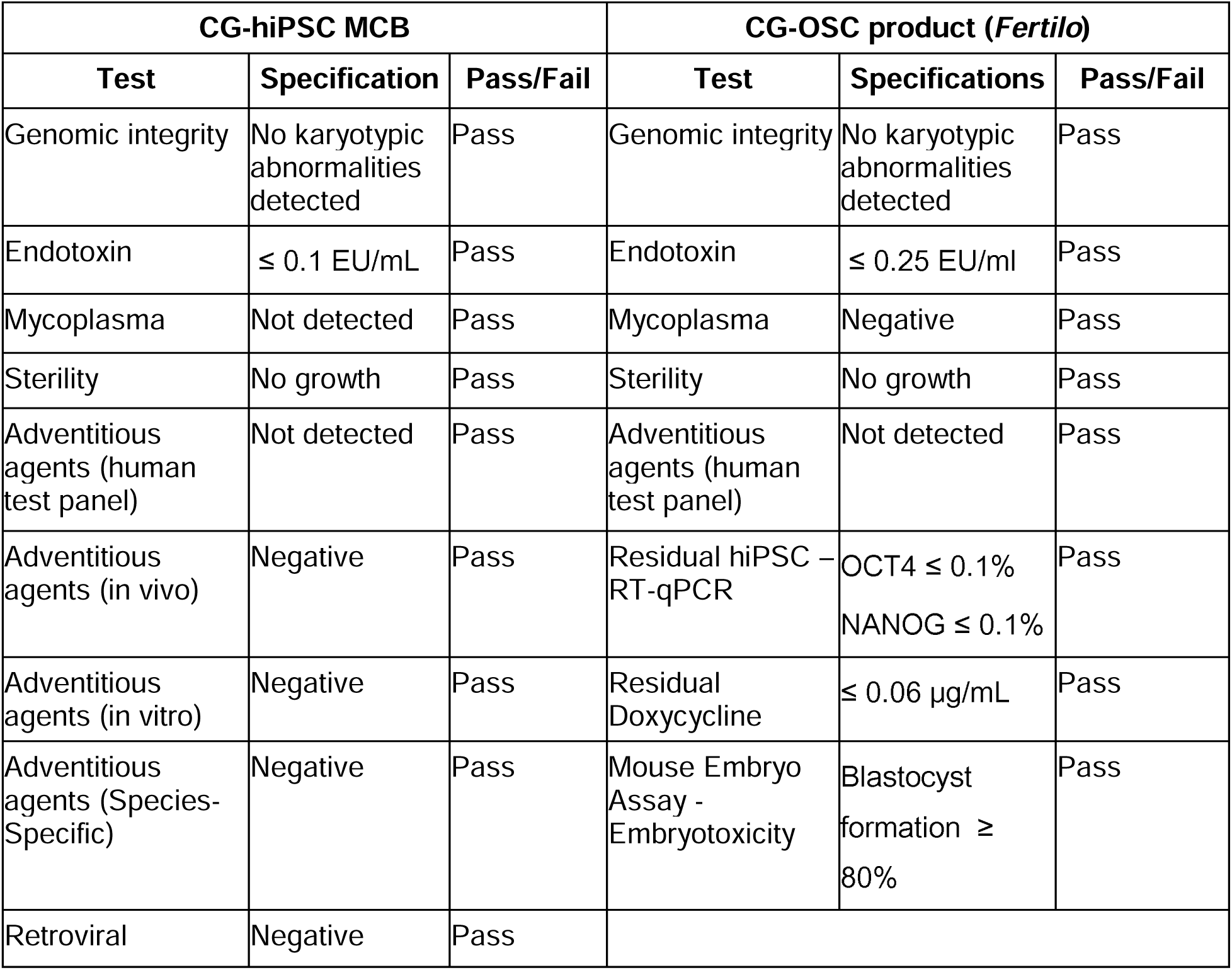
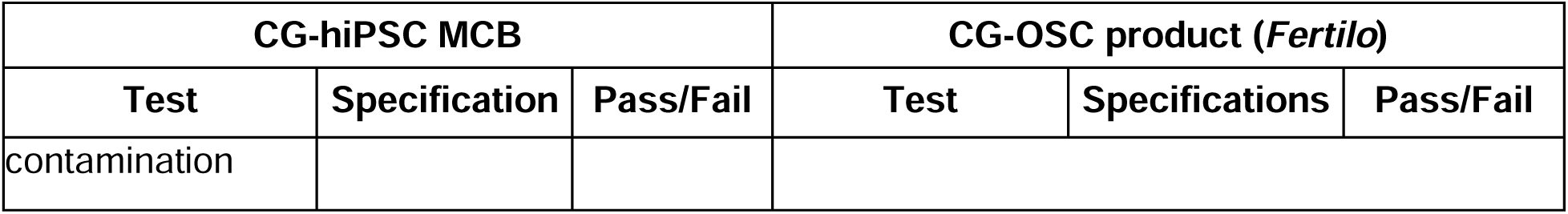
Analytical Testing of the CG-hiPSC MCB and CG-OSC product (*Fertilo*)

Moreover, maintenance of a well-documented chain of custody is critical for ensuring the integrity, traceability, and compliance of the CG-hiPSC to CG-OSC manufacturing process. From the initial generation of CG-hiPSCs in controlled non-classified conditions, through MCB establishment and subsequent CG-OSC production under GMP conditions, stringent documentation and tracking procedures were implemented. Following production, the final product was stored under validated conditions, ensuring stability and quality retention throughout its shelf life. Shipment for distribution happened under temperature-controlled and monitored conditions, thereby mitigating the risk of degradation or contamination. Finally, at the clinical site, proper handling and adherence to protocols ensured that the OSCs maintained potency, safety, and local requirements during application. This end-to-end traceability was fundamental to ensure patient safety and the overall success of the process (**fig. S10**).

### Application of CG-OSCs significantly improves IVM outcomes

To evaluate the clinical application and outcomes of *Fertilo* in humans, a first-in-human observational, longitudinal, cohort study was performed; this study enabled investigation of the safety and efficacy of *Fertilo* application. This study was performed in two phases, the first phase consisting of a multi-center, single arm observational evaluation in twenty patients with the primary purpose of safety evaluation and the establishment of clinical success metrics. The second phase of the evaluation was expanded to a limited comparator-controlled cohort comparison of twenty patents with 1:1 randomization between the two arms at oocyte retrieval to provide measures of efficacy and inform future registrational clinical trial designs.

In phase one, 20 infertile patients under the age of 37 with normal ovarian reserve, as indicated by an anti-mullerian hormone (AMH) value of greater than 2 ng/mL, were recruited for a single arm, multicenter observational evaluation to assess safety. Details about patient demographic information and treatment conditions (**table S8**), as well as the workflow of this study (**fig. S11A**), are provided in the Supplementary Materials. *Fertilo* is intended to support the IVM of immature oocytes collected following minimal stimulation cycles. Thus, following the retrieval of cumulus-oocyte complexes (COCs) and IVM, key embryological and clinical outcomes were assessed. The results for each outcome were determined based on the number of samples that proceeded to each step. First, the MII maturation rate was determined to be 69% per COC retrieved based on the presence of a first polar body (PB1) (**Fig 5A**). Between 16 and 18 hours post-intracytoplasmic sperm injection (ICSI), the fertilization rate was 84% per mature MII based on the formation of two pronuclei (**Fig 5A**). The cleavage rate was determined at 3 days post ICSI based on the presence of 2 or more cells, and the total and high quality (HQ) blastocyst formation rates were determined on day 5, 6, and 7 post ICSI based on cavitation and a 3BB or greater score based on the Gardner Scale, respectively. The cleavage rate was 96%, the blastocyst formation rate was 43%, and the HQ blastocyst formation rate was 38% (**Fig 5A**); these percentages were calculated per fertilized embryo. The euploidy rate, which was determined within 7 days post ICSI via preimplantation genetic testing for aneuploidy (PGT-A) analysis, was 65% per blastocyst biopsy (**Fig 5A**). Finally, the biochemical (implantation) and clinical pregnancy rate were determined per embryo transfer. Biochemical pregnancy was assessed at day 10-14 post transfer based on a β-hCG >5 mIU/mL; the rate of successful implantation was 71% (**Fig 5A**). Clinical pregnancy was assessed at a minimum of 5 to 7 weeks post embryo transfer based on the presence of a visible gestational sac with a fetal heartbeat via ultrasound at 7 weeks’ gestation; the rate of clinical pregnancy was 57% (**Fig 5A**). Notably, the first live birth of a healthy singleton female following an OSC-IVM cycle with *Fertilo* occurred at 38.5 weeks. The baby was 3,255 grams at birth, 49.5 cm, and scored a 9/9 on the Apgar scale, demonstrating no congenital abnormalities after a vaginal birth.

**Fig. 5:**
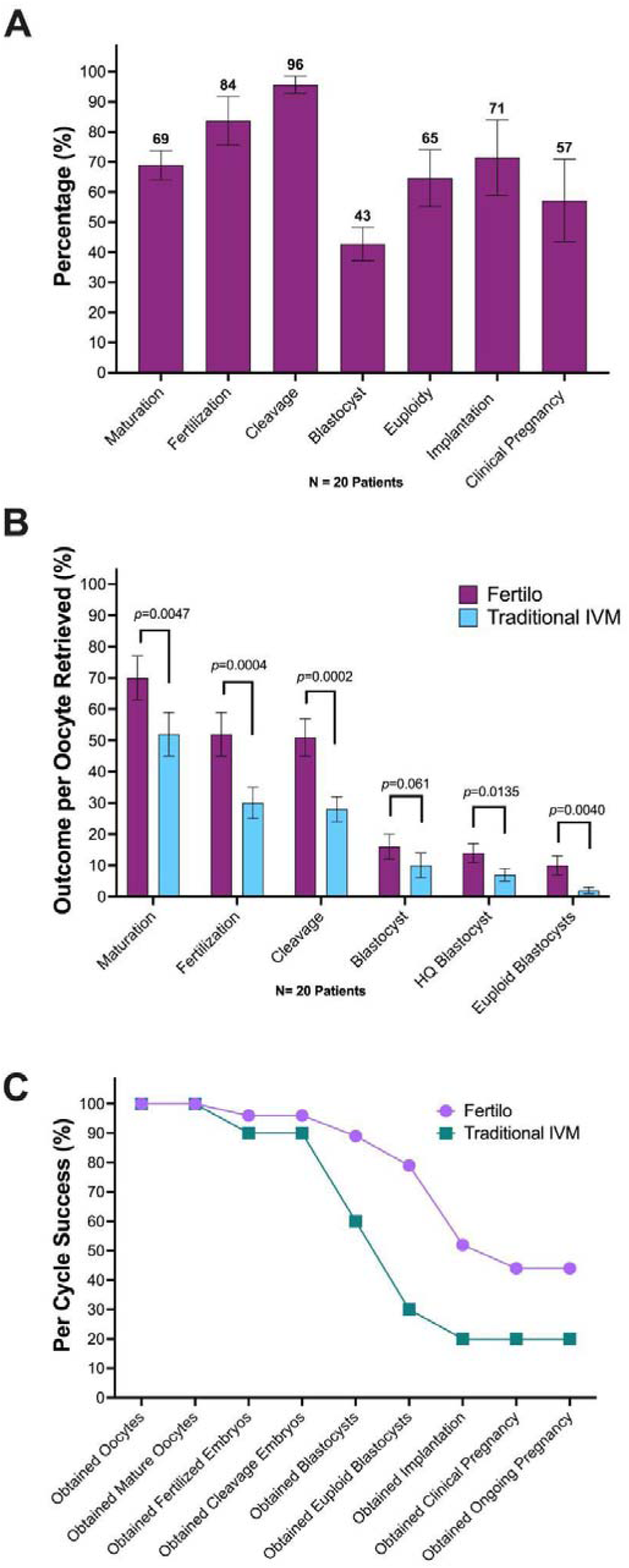
Clinical application of CG-OSCs significantly improves IVM outcomes. A) Clinical outcomes of a single arm, multi-site observational evaluation following OSC co-culture for IVM. The percentages for outcomes were determined incrementally based on the number of samples that proceeded to each step. Data is presented as the mean ± SEM. B) Comparative evaluation of clinical outcomes following IVM co-culture with OSCs (*Fertilo*) versus IVM culture in media alone (Traditional IVM). The outcomes were assessed based on the initial number of oocytes retrieved. Data is presented as the mean ± SEM. C) Compiled results from both clinical evaluations per treatment cycle. Data shows the percentage of treatment cycles that lead to successful completion of each outcome.

After verifying the safety of clinical grade *Fertilo* with a minimal stimulation regimen and collecting measures of success rates, an expansion of this study was initiated. In the second phase, a limited comparator controlled cohort evaluation of *Fertilo* versus traditional IVM (monophasic IVM medium) in a single center, 1:1 randomized evaluation was performed in 20 infertile patients (ten per treatment arm) who were 37 and younger, with an AMH greater than 2 ng/mL. Details about patient demographics and treatment conditions (**table S9**), as well as the workflow of this study (**fig. S11B**), are provided in the Supplementary Materials. The efficacy of *Fertilo* was compared with that of traditional IVM based on embryological endpoints that were analyzed based on the percentage of samples that achieved each outcome per the initial number of COCs retrieved (**Fig. 5B**). Following IVM, the MII oocyte maturation rate was determined; the MII maturation rate of the *Fertilo* group was significantly greater than that of the traditional IVM group (70±7% vs 52±7%, *p*=0.0047; **Fig. 5B**). MII oocytes were fertilized via ICSI, and the fertilization rate per COC was significantly higher in the *Fertilo* group than in the Traditional IVM group (52±7% vs 30±5%, *p*=0.0004; **Fig. 5B**). On day 3 post-ICSI, cleavage was assessed based on the presence of 2 or more cells, and the percent cleavage in the *Fertilo* group was significantly higher than that in the Traditional IVM group (51±6% vs 28±4%, *p*=0.0002; **Fig. 5B**). The overall blastocyst, HQ blastocyst, and euploid blastocyst formation rates were determined within 7 days post ICSI. The rate of blastocyst formation in the *Fertilo* group was slightly but not significantly higher than that in the Traditional IVM group (*p*=0.061; **Fig. 5B**). However, when considering only HQ blastocysts, defined as those with a score of 3BB or greater based on the Gardner Scale, the *Fertilo* group had a significantly higher HQ blastocyst formation rate than the Traditional IVM group (14±3% vs 7±2%, *p*=0.0135; **Fig. 5B**). Finally, the euploid blastocyst formation rate, based on PGT-A analysis, was determined. The euploid blastocyst formation rate of the *Fertilo* group was significantly greater than that of the Traditional IVM group (10±3% vs 2±1%, *p*=0.004; **Fig. 5B**). Notably, 8 of 10 patients in the *Fertilo* group versus only 3 of 10 patients in the Traditional IVM group had at least one euploid blastocyst available for transfer.

Collected data on adverse events up to this point are provided in the Supplementary Materials (**fig. S12**). Adverse events related to minimal stimulation were considered non-*Fertilo* related, as *Fertilo* application in all cases consisted of *in vitro* maturation after oocyte retrieval. Visualizing the compiled data of the full study (both the observational safety phase and expanded comparator-controlled phase) based on successful outcomes per cycle demonstrated that OSC-IVM (*Fertilo*, 44%) has a better treatment success rate than traditional IVM media alone (Traditional IVM, 20%) (**Fig. 7C**); the success rate of treatment is based on the rate of ongoing pregnancy, defined as a fetal sac visualized with ultrasound with detectable fetal heart rate at 10-12 weeks’ gestation. In addition, although the blastocyst and euploid blastocyst rates were the most similar in the outcome comparison per COC, comparison of these rates per cycle shows that *Fertilo* led to substantially more successful blastocyst and euploid blastocyst formation per cycle than Traditional IVM (blastocyst 89% vs 60%; euploid blastocyst 79% vs 30%) (**Fig. 7C**).

## Discussion

Research on female reproductive health has been hindered by the lack of investment, leading to a consequent lack of innovation and new solutions in the field. Recently, it was demonstrated that supplementation of standard of care IVM media with OSCs leads to higher oocyte maturation and euploid embryo formation rates (*11*), possibly due to the mechanisms employed by OSCs to recapitulate the ovarian follicle environment *in vitro*. Here, we demonstrate the process development and clinical application of an hiPSC-derived OSC product known as *Fertilo*. The previously described process for OSC generation (*8*) was optimized using the highest quality raw materials available, and the cellular outcomes and OSC functionality were shown to be conserved after process development. This reproducible process was then applied to generate several clinical OSC lots, which were then subjected to analytical testing and potency assessment via a murine oocyte maturation assay. After the clinical OSC lots were determined fit for release, *Fertilo* was applied via OSC-IVM in a two-phase evaluation with a single arm observational safety cohort phase and a two-arm comparator control cohort phase. Results of the evaluation demonstrated that application of OSCs improves the IVM of human oocytes, leading to favorable embryological and pregnancy outcomes with no *Fertilo*-related adverse events.

Applying the directed differentiation approach to drive granulosa-cell fate is crucial for achieving OSC identity efficiently and seamlessly. As previously reported by our group (*8*), combination of the *NR5A1*, *RUNX2*, and *GATA4* TFs is among the most effective approaches to induce differentiation into OSCs, requiring only minimal supplementation with small molecules without the need for lengthy periods in culture. In this study, this method was utilized to direct the differentiation of a previously described engineered RUO-hiPSC line (*8*) into OSCs, which were capable of improving the rate of MII maturation of human oocytes across multiple differentiation lots. We also demonstrated that the OSCs applied *in vitro* are effectively eliminated post application using NGS-based analysis of *Fertilo* treated human oocytes and subsequently derived embryos, posing minimal risk of carryover or *in vivo* transfer alongside embryos, an important safety consideration.

Raw material risk assessment and sourcing for alternative higher quality reagents contribute not only to reducing the risk of carrying over adventitious agents or toxins to the final product but also to increasing reproducibility from lot-to-lot. Thus, to allow for translation of this technology for clinical applications, several raw materials were substituted with higher quality alternatives, including the differentiation substrate and hiPSC starting material. Due to the complexity and variability of Matrigel, two alternative substrates, vitronectin and laminin-521, were evaluated for OSC differentiation. While both supported successful differentiation, laminin-521 yielded higher cell viability and an improved OSC:hiPSC ratio, making it the preferred substrate moving forward. Subsequently, a donor hiPSC line was engineered using the same strategy as the RUO-hiPSC line (*8*). Nine candidate clones were evaluated, with clone 2-D10 selected as the most suitable line. Notably, despite raw material substitutions and the utilization of a cell line from a different donor, this TF-directed differentiation process consistently generated functional OSCs, demonstrating the robustness of this process.

The substitution of higher quality raw materials also led to a population of OSCs with lower batch to batch variability and a higher percentage of target granulosa-like cells (‘GC’), expressing markers such as *MDK*, *CDH2*, and *ITGB1*. Of note, scRNA-seq results demonstrated strong expression of Midkine (*MDK*) across OSC clusters, as well as high levels of expression of *BMP*, *CYP19A*, and *EGFR*, indicating a predominantly mural granulosa-like cell identity as opposed to a cumulus granulosa-like cell identity (*12, 29–31*). This result supports our previous findings, which also demonstrated significantly upregulated expression of *CYP19A1*— which is essential for estrogen biosynthesis—in response to FSH challenge, a hallmark of mural granulosa cell biology (*12*). These results suggest that the mechanism of action likely relies on recapitulating the ovarian follicle environment *in vitro*, including paracrine signaling from OSCs. Furthermore, previous studies demonstrated that not only are oocytes modulated by OSCs, but the OSC molecular signature is also modulated by the presence of oocytes (*12*), highlighting the importance of employing a dynamic co-culture system for IVM rather than utilization of a static conditioned-media alternative.

A key consideration for clinical development is process consistency; therefore, analytical development was required to assess potency, identity, purity, and safety, ensuring alignment with criteria outlined in the QTPP. As OSCs are derived from hiPSCs, extra safety considerations, including residual hiPSC presence, the detection of communicable diseases, and screening for disease-related agents, are required in addition to the standard attributes for IVF products, such as sterility, endotoxin testing, and embryotoxicity. We demonstrated that multiple production lots consistently met analytical release criteria for key attributes, confirming the efficiency and safety of the manufactured clinical product.

A novel potency assay, referred to as the MOMA, was developed specifically to evaluate the efficacy of different lots of *Fertilo*. This assay evaluates oocyte maturation followed by embryo formation in a surrogate species, mice (*32*), to provide information about the therapeutic activity of hiPSC-derived OSCs. MOMA results showed that OSC-IVM improved the blastocyst formation rate of murine oocytes compared to all negative conditions, demonstrating that this assay is a suitable strategy for OSC batch release.

Following establishment of a clinically-suitable cell product, *Fertilo* was evaluated in a two phase longitudinal cohort analysis: phase I consisted of a single arm multi-center observational safety study and phase II consisted of an expanded comparative evaluation against traditional IVM. To date, these clinical studies have led to 15 ongoing pregnancies, 13 of 30 from *Fertilo* application and 2 of 10 from traditional IVM application. Additionally, *Fertilo* application has resulted in the successful birth of one healthy singleton female. The successful clinical pregnancy outcomes, as well as the first live birth, provides evidence of efficacy and helps establish the favorable safety profile of *Fertilo* and the OSC-IVM method broadly. In the comparative evaluation, *Fertilo* significantly improved the rate of successful embryological outcomes, including MII maturation and euploid blastocyst formation, compared to traditional IVM. Overall, 8 of 10 patients in the *Fertilo* group versus only 2 of 10 patients in the traditional IVM group obtained at least one euploid blastocyst after treatment and proceeded to embryo transfer, suggesting that *Fertilo* improves the rate of successful clinical outcomes based on transferable blastocyst formation rates. In both phases of the evaluation, safety and efficacy analysis of fetal developmental and maternal health remain ongoing, with no reported *Fertilo*-related adverse events in either phase.

Finally, evaluation of the compiled data from the entire study per cycle revealed that *Fertilo* had an overall treatment success rate, defined as ongoing pregnancy, of 44% per cycle, compared to the 20% per cycle for traditional IVM. This result demonstrates that co-culture with CG-OSCs increases the likelihood of successful IVM treatment cycles. Furthermore, previous clinical studies of traditional IVM show ongoing pregnancy rates of approximately 25% per cycle, further confirming that the rate of ongoing pregnancy with *Fertilo* is greater than that seen with standard of care IVM (*14, 15*). Taken together, these data demonstrate the safety and potency of CG-OSCs in promoting successful oocyte maturation, blastocyst formation, and pregnancy following OSC-IVM.

This study represents a significant advancement in women’s health through the clinical development of an hiPSC-derived OSC product; however, delineating certain aspects of OSC-IVM, such as the precise mechanism of action, will still require further investigation. Although gene expression and proteomics analyses support a paracrine mechanism involving crosstalk between OSCs and oocytes, given the ovarian environment’s complexity, future studies with larger cohorts will be required to refine our understanding. Moreover, despite the sample size challenges associated with the utilization of patient-derived samples, we have completed a considerable number of successful embryo transfers. In addition, all patients continue to be monitored for downstream clinical outcomes (biochemical pregnancy, ongoing pregnancy, and live birth), which will provide additional data to evaluate pregnancy endpoints. Notably, the clinical grade, OSC product known as *Fertilo* also recently became the first hiPSC-derived product to obtain investigational new drug (IND) clearance to proceed to Phase 3 clinical trials in the US; these trials will provide crucial potency and safety datasets to further demonstrate the promise of *Fertilo* use. Overall, we demonstrated the first-time clinical development and application of an hiPSC-derived product to advance women’s health. In addition to the application of OSC-IVM, OSCs can serve as a tool with the potential to be further expanded into multiple applications and indications focused on women’s health and infertility. Beyond its current application in OSC-IVM, *Fertilo* could have potential uses in patients with medical conditions that limit the use of hormonal stimulation, such as autoimmune diseases or estrogen-sensitive disorders. Additionally, its ability to support oocyte maturation *in vitro* may provide new fertility preservation strategies for patients undergoing gonadotoxic treatments. Future studies should explore these applications further to expand the clinical scope of OSC-based reproductive therapies.

## Acknowledgement

This work was performed with the support of clinical partnerships at Spring Fertility New York, Extend Fertility, Ruber Clinic of Madrid, Tambre Clinic of Madrid, Pranor Clinic of Lima, and Fertilidad Integral of Mexico City. We thank the dedicated support and work of the embryology and support staff teams at these clinics for coordinating and managing the collaborative study, as well as the technical support of Kathy Potts, Graham Rockwell, Alexa Giovannini, Christopher Collado and Karina Flores. We additionally thank the Wyss Institute for Biologically Inspired Engineering at Harvard University for material transfer of reagents used in the preclinical sections of this work. We thank Professor Mary Herbert, Professor Phillip Jordan, Professor George Church, Professor Kristin Baldwin, Professor Pranam Chatterjee, Dr. Jamie Knopman, and Dr. Sara Vaughn, for advice and guidance on the use of OSCs in IVM work. We also thank the New York University flow cytometry and imaging cores, the Proteomics and Metabolomics Core Facility at Weill Cornell Medicine, Embryotools of Barcelona, and Azenta/Genewiz for their assistance in data generation and analysis.

## Author Contribution

C.C.K., B.P., F.B., A.D.N, and S.P. conceived the experiments. S.P., M.M., I.G.G., E.M.M., E.I.L., P.R., C.C., J.M., W.M., P.P., and L.G., performed all embryology work for the study, C.C.K. supervised their work. B.P., M.J., A.B.F., A.D.N., and F.B. produced and qualified OSC batches, C.C.K. supervised their work. B.P. and S.K. performed bulk- and scRNA-seq analysis and cell type assignments. F.B. performed proteomic experiments and analysis as well as development of the MOMA assay. A.B.F. coordinated the assay logistics and quality process development. C.R., E.N., A.E., L.N.P., J.G., E.M., and P.V., performed on oocyte retrievals and patient treatments. D.R., G.H., P.B., M.D.V., D.A., L.G., P.V., E.M., J.G., and C.C.K. developed and refined clinical treatment protocols and study design and reviewed clinical safety and efficacy outcomes as part of the safety monitoring board, C.C.K., B.P., F.B., and C.L. wrote the manuscript with significant input from all authors.

## Materials and Methods

### Subject ages, ethics, and informed consent

This study was performed according to the ethical guidelines outlined in the Declaration of Helsinki. Oocyte donor participants were enrolled in the study at several fertility clinics using informed consent for donation of gametes for research purposes, with ethical approval from CNRHA 47/428973.9/22 (Spain), Western IRB No. 20225832 (USA), and Protocol No. GC-MSP-01 (Peru), respectively. These study centers performed the non-clinical research associated with oocyte maturation and/or embryo formation endpoints with no subsequent embryo transfer.

For the MOMA, all animal care and procedures were conducted according to protocols approved (reference number 10133) by the Institutional Animal Care and Use Committee of Barcelona Science Park de Barcelona (IACUC-PCB), Spain. The PCB Animal Facility is accredited and registered by Generalitat of Catalonia government (B-9900044) as a breeding and user center for laboratory animal research. All protocols carried out in the PCB Animal Facility comply with standard ethical regulations and meet quality and experimental requirements of current applicable National and European legislation (RD 53/2013 Council Directive; 2010/63/UE; Order 214/1997/GC).

For the assessment involving 40 patients treated as part of a two-phase clinical evaluation, patients were enrolled under informed consent at two fertility centers. Protocol *Fertilo* P1, approved by the ethics committee (CIEI-FMH-USMP, FWA No. 00015320, IRB No. 00003251) was utilized. This evaluation was prospectively registered before recruitment began ISRCTN36032472, with regular IRB updates throughout the two phases as safety and efficacy data became available, to guide treatment in the First in Human evaluation. The protocol, standard operating procedures, and design ensured rigorous oversight and monitoring and compliance with international ethical guidelines.

### Source material

The RUO-hiPSC line was sourced from the laboratory of G. Church; this line is referred to as the F66 line in the reference text (*8*). The RUO-hiPSC line was derived from the NIA Aging Cell Repository fibroblast line AG07141 using Epi5 footprint-free episomal reprogramming. A female donor hiPSC line (VCT-37-F35) was sourced from Reprocell USA (9000 Virginia Manor Rd #207, Beltsville, MD 20705) to serve as the starting material for our CG cell line. The parental CG hiPSC starting material was derived from fibroblasts from the skin punch of an eligible United States donor, ID: RPC-VCT-37. Fibroblasts were isolated from a skin punch biopsy and expanded at Reprocell USA (9000 Virginia Manor Rd #207, Beltsville, MD 20705). Donor identity information for 16 loci was obtained by STR analysis performed on donor blood and fibroblasts to confirm sample identity prior to release of the fibroblasts. Fibroblasts were harvested and cryopreserved in cryopreservation containers (ThermoFisher Scientific, catalog number: 377267) in vapor-phased liquid nitrogen (−196°C) to enable testing and further manufacture. The hiPSC line was generated under GMP conditions using a non-integrative, mRNA-based reprogramming technology. The unmodified parental hiPSC line (VCT-37-F35) was characterized by sterility, endotoxin, STR analysis, Karyotype analysis, mycoplasma, viability, phenotype marker, cell purity and morphology, and differentiation testing.

Donor eligibility and cell line manufacturing were performed in line with relevant regulatory standards and guidelines. Regarding specific stages in the manufacturing process, proper controls were implemented for fibroblast derivation according to established guidelines, while reprogramming and cell expansion took place under fully GMP conditions in compliance with regulatory standards and guidelines of the FDA, EMA, and PMDA. Donor eligibility was determined to be in accordance with 21 CFR Part 1271 Subpart C and FDA Guidance for Industry: Eligibility Determination for Donors of Human Cells, Tissues, and Cellular and Tissue Based Products (HCT/Ps), 2021. All experiments involving human cells were performed according to ISSCR 2021 guidelines (International Society for Stem Cell Research) and approved by the IRB committees.

### Plasmid manufacturing

Plasmids utilized for engineering were manufactured as previously described (*8*). Briefly, transcription factor cDNAs (*NR5A1*, *RUNX2*, and *GATA4*) were obtained as Gateway entry clones from the TFome (*33*) or the ORFeome (*34*). These sequences were cloned into a barcoded Dox-inducible expression vector (Addgene #175503) using MegaGate cloning (*35*). The final *NR5A1*, *RUNX2*, and *GATA4* plasmids (pCK530_22_NR5A1, NM_004959.9; pCK530_33_RUNX2, NM_001024630.4; and pCK530_3_GATA4, NM_001308093) were subjected to whole plasmid sequencing via Plasmidsaurus using Oxford Nanopore Technology with custom analysis and annotation. All plasmids were screened for purity and stored at −20°C, while glycerol stocks of transformed bacteria were stored at −80°C.

### Cell engineering

The parental GMP hiPSC clone VCT-37-F35 was thawed at 37°C and resuspended in NutriStem-XF media (Satorius, 05-100-1A) supplemented with Y-27632 (STEMCELL Tech, 72304). Cells were centrifuged and the supernatant was removed. Then, pelleted cells were again resuspended in NutriStem-XF media (Satorius, 05-100-1A) supplemented with Y-27632 (STEMCELL Tech, 72304) and cell count and viability were assessed. Cells were then seeded onto flasks pre-coated with iMatrix-511 (Nippi, 892005) and incubated for 6 days at 37°C. To adapt hiPSCs to the media and substrate, cells were cultured for 3 passages, during which cell quality was monitored based on morphology and confluency; for each round of expansion, Accutase (STEMCELL Tech, 7922) was used to facilitate cell detachment from flasks, and cell count and viability were assessed. Parental hiPSCs (VCT-37-F35) at passage 11 (day 16) were assessed for confluency, detached using Accutase (STEMCELL Tech, 7922), centrifuged, resuspended in TeSR-AOF supplemented with Y-27672 (STEMCELL Tech, 100-0403), and cell count and viability were assessed. Resuspended cells were centrifuged and resuspended in electroporation buffer (P3 solution, P3 Lonza Nucleofection Kit, V4XP-3024) with equimolar quantities of the 3 plasmids encoding the transgenes *NR5A1*, *RUNX2*, and *GATA4* (pCK530_22_NR5A1, pCK530_33_RUNX2, and pCK530_3_GATA4) in addition to the PiggyBac transposase plasmid (pAMP470 pBASE-Super PiggyBac Transposase). Delivery of the plasmids into the cells was performed using the Lonza 4D-Nucleofector Unit set to the CM-113 program. Transfected cells were plated (100,000 cells/cm^2^) into a well of a 24-well plate pre-coated with Vitronectin-XF (STEMCELL Tech, 7180) and cultured with TeSR-AOF supplemented with Y-23672 (STEMCELL Tech, 100-0403). For the next four days (days 17-20), daily media changes with TeSR-AOF (STEMCELL Tech, 100-0402) supplemented with Puromycin (Sigma-Millipore, P9620-10ML) were performed for selection; cell quality was monitored by morphological and confluency checks. The following four days (days 20-24), resistant cells were maintained with TeSR-AOF with no supplementation (STEMCELL Tech, 100-0402). On day 24, expanded resistant cells were detached from the flasks using Accutase (STEMCELL Tech, 7922), centrifuged, and resuspended into TeSR-AOF supplemented with Y-23672 (STEMCELL Tech, 100-0403). The cells from one well of the 24-well plate (1.9 cm^2^) were further expanded into two wells of a 6-well plate (19.2 cm^2^). From days 25-27, daily media changes with TeSR-AOF (STEMCELL Tech, 100-0402) supplemented with Puromycin (Sigma-Millipore, P9620-10ML) were conducted, and cell quality was monitored by morphological and confluency checks. On day 28, clonal selection was performed by limiting dilution of cells on TeSR-AOF (STEMCELL Tech, 100-0402) supplemented with CloneR^TM^2 (STEMCELL Tech, 100-0691). Single cells were plated onto Vitronectin-XF (STEMCELL Tech, 7180). During clonal expansion (days 29-34), individual wells were carefully monitored to assess colony morphology, growth/confluency, and confirm the presence of a single colony per well. TeSR-AOF (STEMCELL Tech, 100-0402) media changes were performed daily until ready for colony selection. Between days 35-46, once a clone had reached significant levels of expansion (< 10 days after seeding), the individual well was treated with Accutase (STEMCELL Tech, 7922) to promote cell detachment. Cells were centrifuged, resuspended in TeSR-AOF supplemented with Y-23672 (STEMCELL Tech, 100-0403), and seeded onto one well of a 24-well plate pre-coated with Vitronectin-XF (STEMCELL Tech, 7180). When a given clone was ready to be passed, the same procedure was repeated and expanded cells were plated onto a 6-well plate pre-coated with Vitronectin-XF (STEMCELL Tech, 7180). During this expansion, all clones were screened for integration of the three transcription factors (*NR5A1*, *RUNX2*, and *GATA4*). In addition, TF insertion sites were verified by Whole Genome Sequencing (Azenta). The analysis showed no evidence of mutations in 20 common proto-oncogenes. Expanded cells were treated with Accutase (STEMCELL Tech, 7922), centrifuged, and the supernatant was removed. Cells from each clone were resuspended in CryoStor® CS10 (STEMCELL Tech, 7930), gently mixed, and filled into cryovials. The finished vials were cryopreserved in a Mr. FrostyTM freezing container at −80°C freezer for a maximum of 24 hours. The cryopreserved cells were stored in cryopreservation containers (Azenta Life Sciences, catalog number: 67-0755-11) and transferred to vapor-phase liquid nitrogen (−196°C) with temperature monitoring for long-term storage.

### Cell screening, selection, and preliminary characterization

After seed clones were assessed by genotyping PCR to verify the presence of the three TFs, nine clones harboring all the TFs were identified and each clone was subjected to a more in-depth screening process. To identify the lead candidate, each of the nine clones was individually differentiated and subjected to a series of assays to ensure identity (pluripotency markers, and genotyping), conformance (cell count and viability), and potency (OSC production and function). The leading candidate clone (2-D10) selected to be used as the starting material for the CG cell line was named CG-hiPSC.

### hiPSC maintenance and OSC differentiation

hiPSCs were maintained in feeder-free conditions and cell culture plates were pre-coated with either Matrigel (Corning), Laminin-521 (StemCell Technologies), or Laminin-511 (Reprocell). Cells were maintained in either mTESR1 (StemCell Technologies) or TeSR-AOF (StemCell Technologies) without antibiotics at 37 °C in 5% CO_2_. All hiPSCs were verified for expression of markers associated with pluripotency (TRA-1-60 and TRA-1-81), negative for mycoplasma or other human adventitious agents (performed by IDEXX Bioanalytics), and karyotypically normal (G-band karyotype test, performed by WiCell Research Institute, and Karyostat, performed by ThermoFisher). RUO-hiPSCs and CG-hiPSCs were authenticated by SNP array (CellID, performed by ThermoFisher). All OSC differentiations were initiated from previously characterized hiPSC working banks under passage 25. Characterization of hiPSC working cell banks includes assessment of marker expression, morphology, vial appearance, container closure integrity, cell count and viability post-thaw, differentiation potential, and testing for endotoxin, mycoplasma, sterility, adventitious agents, and genomic integrity. OSC differentiation was performed as previously described (*8*). In short, hiPSCs were exposed to the Rho kinase inhibitor, Y-27632, and the WNT activator, CHIR99021, to prime the cells into a mesodermal fate. Exposure to doxycycline throughout the process induced overexpression of the TFs *NR5A1*, *RUNX2*, and *GATA4*. The differentiation process required five days in culture. All images were taken with an ECHO Revolve Microscope (Discover ECHO).

### Cell count and viability

Cell counts and viability assessment was performed using an Eve Automated Cell Counter (NanoEnTeck) and a NucleoCounter NC-202 (ChemoMetec).

### Single cell RNA sequencing (scRNA-seq)

OSCs were cryopreserved in CryoStor CS10 (StemCell Technologies) prior to being processed for single-cell RNA sequencing by Genewiz (Azenta). Cells were loaded onto a Chromium Single Cell Chip (10x Genomics) and processed through the Chromium Controller to generate single-cell gel beads in emulsion. scRNA-seq libraries were generated using the Chromium Single Cell 3′ Library & Gel Bead Kit (10x Genomics). Target cell recovery was estimated to be 3,000 cells per sample. Files were generated following a split-set analysis workflow. First bcl2fastq was used to generate the fastq files. Next, a reference genome was generated using split-pipe and the Homo sapiens GRCh38 file, also using STAR (*36*) and Samtools (*37*). Then, split-pipe was run to process and align the files against the reference genome. The resulting files were generated in an mtx matrix format. Finally, the files were combined, and an Anndata object was made in h5ad file format. For the analysis, cells with less than 200 genes were filtered out, as well as genes found in less than 3 cells. Cell counts were normalized to 10,000 Unique molecular identifiers (UMIs) per sample and log (ln) plus 1 transformed. Principal component analysis was performed using the Scanpy package (v1.9.6) based on 30 PCA components, and using PCA results, nearest neighbor analysis was performed using n_neighbors = 20. Batch correction was performed using Scanpy’s ComBat method (*38*). The number of components used for batch correction was 30 and the data was then transformed using the Uniform Manifold Approximation and Projection (UMAP) method (*39*). Clusters were formed using the Leiden method (*40*) with a resolution of 0.25 for the complete dataset (containing all lots described in the manuscript), and projected into the subsetted objects. At first, 15 Leiden clusters were identified. Thereafter, clusters were combined based on biological similarities, which resulted in nine final clusters: Early GC I, Early GC II, Early GC III, GC I, GC II, GC III, Atresia/luteolysis, Mitochondrial gene enriched (Atresia/luteolysis), and Ribosomal gene enriched (Atresia/luteolysis). The marker genes per cluster, as well as certain granulosa cell markers (*GJA1, MDK, BBX, HES4, PBX3, YBX3, BMPR2, CD46, COL4A1, COL4A2, LAMC1, ITGAV,* and *ITGB1*), were analyzed to identify cluster cell types. Dot plots and feature plots were generated to assess the expression levels of certain genes per cluster or per sample. Based on downstream analysis, clusters 0 and 5 were subsetted from the original object and they were re-clustered using the Leiden method at a higher resolution of 0.3. Subclusters were combined based on biological similarities. Once all the subgroups were identified, the subclusters resulting from cluster 0 and cluster 5 were merged with the original object. Gene signatures based on genes in the folliculogenesis stage (*18*) were also analyzed and used in predicting cluster identification. Signature scores for the RUO-OSC-M subset were generated for two groups: Antral and Pre-Ovulatory genes. Based on the final object, 4 more subsets were generated: (1) a RUO-OSC-M object consisting of the following lots: lot 6, lot 7, lot 8, lot 29, lot 48, and lot 56; (2) a RUO-OSC-L object consisting of lot 77 and lot 86; and (3) a CG-OSC-L object consisting of lot 88 and lot 90. UMAPs of all the subsets were generated with the cluster names from the original object.

### Bulk RNA-sequencing

Libraries for RNA sequencing were generated using the NEBNext Ultra II Directional RNA Library Prep Kit for Illumina (NEB #7765L) in conjunction with NEBNext Multiplex Oligos for Illumina (Unique Dual Index UMI Adaptors RNA Set1, NEB #7416S) and NEBNext Poly(A) mRNA Magnetic Isolation Module (NEB #E7490L), according to the manufacturer’s instructions. The library pool was sequenced at Azenta using Illumina 2×150 bp, ~350 M PE reads (~105 GB), lightening package. Illumina sequencing files (bcl-files) were converted into fastq read files using Illumina bcl2fastq (v2.20) software deployed through BaseSpace using standard parameters. RNA-seq data gene transcript counts were aligned to *Homo sapiens* GRCH38 (v2.7.4a) genome using STAR (v2.7.10a) (*36*) to generate gene count files and annotated using ENSEMBL (*36*). Gene counts were combined into sample gene matrix files (h5). Computational analysis was performed using the Scanpy (v1.9.6) package. Two h5ad files were joined based on similar features and genes. The two merged files were created into one Anndata object which was normalized to 10,000 UMI per sample and log (ln) plus 1 transformed. Principal component analysis was performed using 30 PCA components. Projection into two dimensions was performed using the UMAP method (*39*). This analysis was performed to evaluate the CG-hiPSC sub-clones: 2-A7, 2-C9, 2-D10, 2-G1, 2-G11, 2-H10, 3-C7, 3-D3, and 3-E3.

### Barcode-Seq identification of residual OSC presence in human oocytes and embryos

Low input bulk RNA sequencing of individual post-IVM-oocytes was performed using the NEB Low Input RNA-seq library preparation kit and sequenced on an Illumina MiSeq. Two reference positive control samples of RUO-OSCs were directly sequenced to serve as controls, using the same library preparation and sequencing kit. Additionally, two negative control samples of patient derived cumulus cells were used as a somatic cell negative control and likewise prepared using the same library preparation and sequencing kit. All samples were assessed for RNA integrity scores (RIN) of greater than 8 and sequenced to the same depth.

In order to determine the residual presence of OSCs using RNA-sequencing, a set of sequences unique to OSCs was utilized as a target for assessment. This unique sequence consists of a 20 base pair barcode that is expressed uniquely on the 3’ untranslated region (UTR) or the exogenous transcription factors (TF) used to manufacture *Fertilo* OSCs.

Barcode identification was performed on Fastq files using the following method. Briefly, a Kallisto reference genome library was built using Kallisto index, depending on target sequences, using kmers = 15 to kmers =31. An index on specific barcode sequences and flanking regions down stream of inserted TF sequences was included. Additionally, an index reference genes (KRT19 and EpCAM) known to be active in OSC lines and oocyte cells was included. Kallisto sudo alignment of transcriptomic read files (FASTQ) to kallisto genomic index was performed. Read counts were then collected and compared of barcode sequences found down stream of inserted synthetic transcription factor (TF) genes.

To ensure the oocytes were sequenced with enough depth and were not negative due to low read depth, two highly expressed housekeeping genes were likewise analyzed, KRT19 and EpCAM, to filter samples with unsuitably low read depth. These housekeeping genes were chosen for their conserved expression in both oocytes and reference somatic cell and *Fertilo* OSC controls. Relative quantification of the barcodes and gene sequences of interest were performed using kallisto pseudoalignment discovery (v 0.44.0) and expressed as a transcript per million (TPM) measure.

To select barcodes that are sensitive and specific to detection of *Fertilo* OSCs, three 20 bp barcodes, with 2 kmer lengths, as well as the endogenous gene were evaluated for sensitivity and specificity. Sensitivity was assessed as being detectable in the reference RUO OSC positive control sample. Specificity was assessed as being detectable in the reference RUO OSC positive control sample and undetectable in the cumulus cell somatic negative control.

For analysis of residual *Fertilo* OSCs in embryo samples, the preimplantation genetic testing for aneuploidy (PGT-A) of trophectoderm biopsies was utilized. The platform utilized for PGT-A utilized microarray analysis (Juno Genetics) on 4 to 8 cell biopsy samples. The analysis utilized single nucleotide polymorphisms (SNPs) and haploblock mapping to determine whether the biopsy sample is euploid, aneuploid, segmental aneuploid, or mosaic. Furthermore, for each embryo, a sample of maternal blood and paternal sperm were utilized to compare the SNPs of the embryo to the SNPs of the maternal and paternal donors. Furthermore, a sample of RUO OSCs was provided to determine the SNP profile of residual RUO OSCs. Therefore, through analysis of the embryo’s SNPs presence of maternal, paternal, and residual *Fertilo* OSC DNA could be accurately identified in parallel to aneuploid status. For all biopsies, day 3 laser-assisted zona drilling was performed, and biopsies were performed on embryos that reached transferable quality grade on day 5 through 7 of development, as determined by the Gardner Score of 3BB or greater.

### Design of Experiments (DOE)

DOE was conducted using JMP software (JMP 17, SAS Institute Inc., Cary, NC, USA) to optimize experimental parameters, including media supplementation, substrates, and doxycycline treatment designs. Factors were selected from established modulators for OSC specification and factor ranges were determined through literature review (**table S2**). Responses were FOXL2 expression and viability. Viability was used to screen and remove experimental groups below 50% cell survival prior to the final statistical analysis. A custom design was employed and optimized for D-optimality to investigate main effects and interactions. JMP software facilitated generation of the experimental design matrix, as well as statistical analysis through response surface methodology (RSM), ANOVA, and optimization of conditions to maximize response desirability for FOXL2 expression.

### Proteomics

Liquid chromatography followed by tandem mass spectrometry (LC-MS/MS) analysis was conducted on a series of samples, including RUO-hiPSC (n=1), CG-hiPSC (n=1), RUO-OSC-M lot 56 at time 0h (n=1) and after 24h (n=1) of culture with supplemented IVM media, and CG-OSC-L lot 88 at time 0h (n=1) and after 24h (n=1) of culture with supplemented IVM media. Each condition involved the analysis of 2 million cells. Supplemented IVM media consisted of IVM media (Origio) supplemented with 75 mIU/mL of recombinant FSH (Millipore), 100 mIU/mL recombinant hCG (Sigma), 500 ng/mL of androstenedione (Sigma), 1 ug/mL of doxycycline (StemCell Tech) and 10 mg/mL of human serum albumin (HSA; Life Global). Conditioned media derived from RUO-OSC-M lot 56 (n=1) and CG-OSC-L lot 88 (n=1) were also analyzed. To generate conditioned media, 2 million OSCs were cultured in 2 mL of supplemented IVM media for 24 hours, maintaining the ratio of the intended clinical cell dose of 1,000 OSC cells per 1 µL of media. The 24-hour culture was performed with CO_2_ set for a pH of 7.2-7.4. Following culture, OSCs and conditioned media were separated and processed independently. Supplemented IVM media without OSCs was used as a media control. The conditioned media were subjected to consecutive centrifugations (300 x g, 1,200 x g, and 3,000 x g) to remove cellular remnants, and then passed through albumin depletion columns (AVK-50, AlbuVoid Albumin Depletion Columns Biotech Support Group) to eliminate HSA-derived albumin. Proteins from cells and conditioned media were precipitated using acetone, re-suspended in 0.1% RapiGest and 25 mM ammonium bicarbonate, reduced with DTT, and alkylated with iodoacetamide, before undergoing in-solution trypsin digestion overnight at 37°C. The resulting peptides were desalted using C18 stage-tip columns prior to analysis using a Thermo Fisher Scientific EASY-nLC 1200 coupled online to a Fusion Lumos mass spectrometer (Thermo Fisher Scientific). For LCMS/MS analysis, buffer A (0.1% FA in water) and buffer B (0.1% FA in 80 % ACN) were used as mobile phases for gradient separation. For peptide separation, a packed in-house 75 µm x 15 cm chromatography column (ReproSil-Pur C18-AQ, 3 µm, Dr. Maisch GmbH, German) was used. Peptides were separated with a gradient of 5–40% buffer B over 30 min, and 40%-100% B over 10 min at a flow rate of 400 nL/min. Fusion Lumos mass spectrometer operated in a data independent acquisition (DIA) mode, collecting MS1 scans in the Orbitrap mass analyzer from 350-1400 m/z at 120K resolutions. The instrument was set to select precursors in 45 x 14 m/z wide windows with 1 m/z overlap from 350-975 m/z for HCD fragmentation. MS/MS scans were collected in the orbitrap at 15K resolution. Data analysis involved searching against human Uniprot database (8/7/2021) using DIA-NN v1.8 with filtering for 1% false discovery rate (FDR) for both protein and peptide identifications. Protein intensities were normalized and log transformed for relative quantitation, and multiple hypothesis correction of p-values was performed using the Benjamini-Hochberg method. Proteomic analyses were conducted at the Proteomics and Metabolomics Core Facility at Weill Cornell Medicine (New York, USA). GO Chord graphs generated with the free online platform, SRplot (*41*).

### Immunostaining (immunofluorescence and flow cytometry)

Immunofluorescence staining was conducted on fixed hiPSCs adhered to a slide, following the protocol recommendations from the New York Stem Cell Foundation (NYSCF). Briefly, hiPSCs were fixed in 4% paraformaldehyde (PFA), followed by a blocking step in a blocking buffer (3% donkey normal serum and 0.1% Triton-X). The primary antibodies were mouse monoclonal antibody against OCT3/4 (1:200; sc5279, Santa Cruz Biotechnology), goat polyclonal antibody against SOX2 (1:50; AF2018, R&D systems), goat polyclonal antibody against NANOG (1:50; AF1997, R&D systems), and Alexa Fluor 488 mouse monoclonal antibody against TRA-1-60 (1:100; 560173, BD Biosciences). The secondary antibodies were Alexa Fluor 555 donkey anti-mouse IgG (A32773, Invitrogen), Alexa Fluor 488 donkey anti-goat IgG (A32814, Invitrogen), and Alexa Fluor 647 donkey anti-goat IgG (A32849, Invitrogen). All antibody dilutions were prepared in a blocking buffer and incubated at room temperature (RT) for 1 hour. After incubations, samples underwent three washes of 30 min each with PBS containing 0.1% Tween-20 (PBST). Subsequently, samples were mounted with Prolong Gold mounting medium prior to imaging using an ECHO Revolve microscope.

Flow cytometry analyses were conducted on live and fixed RUO-OSC and CG-OSC samples. For the analysis of live cells, cells were incubated with a PE-conjugated mouse monoclonal antibody against CD82 (1:50 dilution; 342104, BioLegend) in FACS wash (dPBS with 5% fetal bovine serum (FBS)). After incubation, cells were washed with FACS wash, stained with propidium iodide (1:20 dilution; P4864, Millipore Sigma) for live/dead staining, and subsequently analyzed using a CytoFlex Flow Cytometer. For the analysis of fixed cells, cells were fixed with 4% PFA for 15 minutes at RT and then washed with dPBS. After, cells were permeabilized using FACS wash solution containing 0.1% Triton X-100 (A16046.AE, Thermo Fisher Scientific). The primary antibodies were mouse monoclonal antibody against OCT3/4 (1:50 dilution; sc5279, Santa Cruz Biotechnology) and rabbit polyclonal antibody against FOXL2 (1:100 dilution; A16244, ABclonal). The secondary antibodies were Alexa Fluor 555 donkey anti-mouse IgG (A32773, Invitrogen), and Alexa Fluor 488 donkey anti-rabbit IgG (A32790, Invitrogen). Following incubations, cells were washed with FACS wash containing Triton X-100 and then analyzed using a CytoFlex Flow Cytometer. Unstained cells (negative controls) were used to determine the gating strategy.

For analytical testing, hiPSCs were stained with Alexa Fluor 488-conjugated TRA-1-60 antibody (1:100 dilution; TRA-1-60 Monoclonal Antibody Alexa Fluor 488, BD Pharmigen™, Cat.#: 560173) for 1 hour on ice. Subsequently, cells were stained with propidium iodide (1:20 dilution; Sigma, Cat.#: P4864) right before analysis to distinguish live/dead cells. A total of ≥ 30,000 TRA-1-60+ live cells were acquired on a CytoFLEX S Flow Cytometer and the data was analyzed using FlowJo 10.10.0. The gating strategy was defined using an unstained hiPSC sample.

### RT-qPCR and genotyping PCR

For genotyping PCR, DNA extraction from various hiPSC clones was carried out using the QuickExtract DNA Extract Solution (Epicentre), following the manufacturer’s instructions. PCR amplification was performed using Q5 High-Fidelity 2X Master Mix (New England Biolabs) for 35 cycles with a 20-second extension time. Subsequently, PCR was performed to validate the integration of the 3 TFs *NR5A1*, *GATA4*, and *RUNX2*. The PCR protocol involved an initial denaturation step at 98°C for 30 seconds, followed by 35 cycles of denaturation at 98°C for 10 seconds, annealing at 66-70°C (*NR5A1*, 67°C; *RUNX2*, 66°C; *GATA4*, 70°C) for 10 seconds, and extension at 72°C for 20 seconds, with a final extension step of 2 minutes. The reaction was then held at 4°C. Gel electrophoresis (2% agarose) was performed to confirm the presence of TFs.

RT-qPCR was performed to assess the gene expression of *POU5F1* and *NANOG*, following protocol recommendations from the New York Stem Cell Foundation (NYSCF). RNA extraction was performed using the Quick-RNA Microprep Kit (Zymo Research) following the manufacturer’s instructions. cDNA synthesis was carried out with the LunaScript RT SuperMix Kit (New England Biolabs). Quantification of RNA and cDNA was performed using Nanodrop. PowerUp SYBR Green Master Mix (Applied Biosystems) was used for RT-qPCR. For RT-qPCR, the thermocycler program consisted of a primer annealing stage at 25°C for 2 minutes, followed by cDNA synthesis at 55°C for 10 minutes, concluded with heat inactivation at 95°C for 1 minute. The RT-qPCR protocol involved an initial denaturation step at 95°C for 2 minutes, followed by 40 cycles of denaturation at 95°C for 3 seconds, annealing at 60°C for 30 seconds, and an analysis step.

### Functional assessment (oocyte maturation)

The oocyte maturation-stimulating potential of various OSC batches was used to evaluate potency. Briefly, human immature oocytes surrounded by cumulus cells, known as cumulus-oocyte complexes (COCs), were retrieved from subjects that underwent minimal stimulation protocols. Subsequently, these immature COCs were co-cultured with different batches of OSCs for 24-30 hours to facilitate IVM. After IVM, oocytes were evaluated for their maturation state and categorized into immature stages (GV oocyte or MI oocyte) or mature stage (MII oocytes). MII oocyte maturation rate (%) was calculated by dividing the total number of mature MII oocytes by the initial number of immature oocytes; MII oocyte maturation rate was used as the potency readout. For a comprehensive description of the methods follow, refer to our recent publication (*11*). Sibling oocytes were used for initial comparisons described in **fig. S2B**. For all the other comparisons, the control and OSC-IVM groups consist mostly of oocytes from non-overlapping donor cohorts. All analyses were performed using One-way ANOVA (Dunnett’s multiple comparison test). The number of oocyte donor participants and the total number of oocytes used per OSC batch assessed are detailed in **table S7**.

### Control conditions for hiPSC manufacturing

The controlled conditions used for manufacturing were established based on the current Good Manufacturing Practice (cGMP) conditions outlined in 21 CFR Parts 210 and 211 with a focus on use of suitable materials, implementation of contamination and cross contamination controls, and adherence to standard operating procedures. Briefly, a quality control unit was established at Gameto and all personnel and consultants met qualifications for assigned functions; all buildings and facilities were deemed suitable for manufacturing; equipment was determined to be appropriate for use with detailed maintenance and cleaning procedures; raw materials were received, stored, handled, and tested appropriately; standard operating procedures for production and processing were written and executed, and deviations were reported; packaging, labeling, holding, and distribution procedures were put in place; and protocols for salvaged drug products were developed. In addition, suitable laboratory controls, as well as procedures for maintaining records and generating reports, were established.

### Safety testing

Mycoplasma testing was performed on OSCs in duplicate. Mycoplasma testing included detection of the presence of *Mycoplasma pulmonis* and *Mycoplasma sp.* DNA was extracted and real-time quantitative polymerase chain reaction (RT-qPCR) was used for the direct detection of nucleic acid sequences for the mycoplasma targets. In addition, mycoplasma testing was also performed on cell culture supernatant collected during harvesting day. Cell culture supernatant was inoculated into a single flask containing sterile complete mycoplasma broth. The flask was incubated under microaerophilic conditions at 36°C ± 1°C under 5-10% CO_2_ in nitrogen for 7 days. Broth culture was collected, RNA was isolated, and RT-qPCR was then performed to amplify and detect nucleic acid sequences from organisms within the class Mollicutes, including mycoplasma species and related *Acholeplasma laidlawii*.

Endotoxin testing was performed on thawed hiPSC or OSC samples using a quantitative kinetic chromogenic LAL (Limulus Amebocyte Lysate) assay in accordance with the USP <85> compendial method.

Sterility testing was performed in accordance with the USP <71> compendial method to detect bacteria and fungi. Samples are aseptically directly inoculated onto vessels containing Fluid Thioglycollate Medium (FTM) and Soybean Casein Digest Medium (SCDM)/Tryptic Soy Broth (TSB). Two vessels containing uninoculated FTM and two vessels containing uninoculated SCDM/TSB serve as negative controls. FTM vessels are placed at 31°C – 35°C for 14 days, whereas SCDM/TSB vessels are placed at 21°C – 25°C for 14 days. All vessels are observed for evidence of growth on days 3, 4 or 5, day 7 or 8, and day 14. The test is considered valid if no growth is observed in the negative controls.

Adventitious agents (human test panel) testing was performed on hiPSCs and OSCs. Adventitious agents (human test panel) testing included detection of the presence of the most common human adventitious agents: Hepatitis A, Hepatitis B, Hepatitis C, HTLV Type I & II and STLV Type 1, Human Immunodeficiency Virus Type 1, Human Immunodeficiency Virus Type 2, Epstein Barr Virus, Human Cytomegalovirus, Human Herpesvirus Type 6, Human Herpesvirus Type 7, Human Herpesvirus Type 8, Parvovirus B19, JC Virus, and BK Virus. DNA is extracted, and real-time quantitative polymerase chain reaction (qPCR) is used for the direct detection of nucleic acid sequences for Hepatitis B, Epstein Barr Virus, Human Cytomegalovirus, Human Herpesvirus Type 6, Human Herpesvirus Type 7, Human Herpesvirus Type 8, Parvovirus B19, JC Virus, and BK Virus. RNA is extracted, and real-time reverse transcription quantitative polymerase chain reaction (RTqPCR) is used for the direct detection of nucleic acid sequences for Hepatitis A, Hepatitis C, HTLV Type I & II and STLV Type 1, Human Immunodeficiency Virus Type 1, and Human Immunodeficiency Virus Type 2.

Genomic integrity of hiPSCs and OSCs is assessed through whole genome sequencing (WGS). Sample processing is performed using the PacBio Nanobind PanDNA Kit, followed by library preparation with the SMRTbell® Prep Kit 3.0. Sequencing is then conducted on the PacBio Revio, a long-read sequencing system that utilizes single-molecule real-time (SMRT) sequencing technology. To evaluate genome-wide DNA alterations that may have arisen during the manufacturing of hiPSC banks and *Fertilo* OSC lots, variant calling analysis is performed. The effects of detected variants on transcripts are annotated and predicted using Ensembl. Comparisons are made between the test sample DNA and the GRCh38 reference genome, with intersection tests conducted for single nucleotide polymorphisms (SNPs), insertions/deletions (INDELs), and structural variants (SVs).

The presence of residual hiPSCs in the test article (*Fertilo*) is assessed by RT-qPCR targeting the pluripotent stem cell markers *OCT4* and *NANOG*. GAPDH serves as a housekeeping gene for normalization. The acceptance criteria for Residual hiPSC testing is “*OCT4* ≤ 0.1% and *NANOG* ≤ 0.1%”.

Residual doxycycline was performed on OSC samples. Exposure to doxycycline throughout the hiPSC differentiation process induces overexpression of the transcription factors, NR5A1, RUNX2, and GATA4, directing differentiation of hiPSCs towards OSCs. To confirm that doxycycline is not carried over into the final product, an ELISA against doxycycline (Abcam, Cat. #ab285232) was performed on conditioned media.

Embryotoxicity is tested using a mouse embryo assay (MEA) to confirm that *Fertilo* does not cause harm to embryos or gametes, following the FDA guidance recommendations on conducting “Mouse Embryo Assay for Assisted Reproduction Technology Devices”. At the completion of embryo culture, blastocysts formation rate (BFR) is assessed, and the acceptance criteria is ≥ 80% embryos successfully developed into blastocysts.

### Murine oocyte maturation assay (MOMA)

Hybrid (B6/CBA) females, and male mice from the same genetic backgrounds, were purchased from Janvier Labs. Upon arrival, all mice were quarantined and acclimated to the PCB Animal facility (PRAAL) for approximately 1 week prior to use. Mice were housed with a 12-hr light/dark cycle (lights on at 7:00 A.M.) with ad libitum access to food and water. All procedures involving mice, e.g., handling, administration of hormones for superovulation of females were conducted at the PRAAL Animal Facility, while procedures involving oocyte manipulation, e.g., oocyte collection, oocyte insemination and embryo culture up to blastocyst were performed at Embryotools’ laboratories.

A total of 10 mL of *Fertilo* IVM media, consisting of IVM Base Media (Cooper), 10 mg/mL human serum albumin (HSA) (Cooper), 75 mIU/mL recombinant follicle stimulating hormone (rFSH) (Sigma), 100 mUI/mL human chorionic gonadotropin (hCG) (Sigma), and 500 ng/mL androstendione (Sigma), and 10 mL of IVM control media, consisting of IVM Base Media (Cooper), 10 mg/mL HSA (Cooper), 75 mIU/mL rFSH (Sigma), 100 mUI/mL hCG (Sigma), and 500 ng/mL androstendione (Sigma), were prepared and stored at 4°C until use. Plating dishes were prepared 16-24 hours prior to oocyte culture to allow for equilibration. The wells of a universal GPS dishes (UGPS-010, Cooper) were prepared with 50 µL LAG media, 100 µL *Fertilo* IVM media, 100 µL IVM control media, or 50 µL *Fertilo* IVM media/ IVM control media for wash droplets. Then, 12 mL of mineral oil (Kitazato) was used to overlay droplets. All dishes were incubated overnight at 37°C with CO2 set for a pH of 7.2-7.4 and 5% O2.

On the day of oocyte culture, *Fertilo* OSCs were seeded 2-5 hours before co-culture. To thaw *Fertilo* OSCs, one OSC cryovial was removed from liquid nitrogen storage and placed upright into a 37°C water bath or dry bead bath to incubate for 2-3 minutes or until ice dissolved. Then, 400 µL of pre-warmed *Fertilo* IVM media was added to the vial and the cells were mixed by pipetting. After, 500 µL of the cell suspension was added to a 1.5 mL Eppendorf tube and centrifuged at 300 x G for 5 minutes at room temperature (RT). The supernatant was then removed and the cells were gently resuspended in another 400 µL pre-warmed *Fertilo* IVM media. Following centrifugation at 300 x G for 5 minutes at RT, most supernatant was removed while ensuring not to disturb the pellet. The cells were then resuspended in pre-warmed *Fertilo* IVM media to 100,000 OSCs per 50 µL according to batch specifications provided in the *Fertilo* Certificate of Analysis. Then, the plating dishes were removed from the incubator and 50 µL of media was removed from the wells with *Fertilo* IVM media. After gently pipetting the cell suspension, 50 µL of OSCs were added to the wells with *Fertilo* IVM media, for a concentration of 100,000 OSCs in 100 µL media. The plate was then placed back into the 37°C incubator for at least 2 hours until oocyte seeding.

Fresh immature mouse oocytes at the germinal vesicle (GV) stage were collected from the ovaries of hybrid (B6/CBA) females between 6 and 8 weeks of age stimulated with 7.5 IU pregnant mare serum gonadotropin (PMSG) 48 hours before retrieval. GVs with cumulus cells were then randomly split between the *Fertilo* IVM group or the IVM control group. Oocytes were then plated and the plates were incubated at 37°C for 18 hours for IVM. After IVM, oocytes were stripped using hyaluronidase (H3884-50MG, Sigma-Aldrich) and assessed for maturity. Mature oocytes with first polar body extrusion were selected for intracytoplasmic sperm injection (ICSI). Fresh sperm from B6/CBA hybrid strain mice was obtained in a microdroplet of culture medium and cultured for 15 minutes at 37°C, 5% CO2, and 5% O2. After incubation, 3 µL of the concentrated sperm solution was further diluted in a 150 µL droplet of the same culture medium. ICSI was performed using a piezo drill-based protocol optimized for the mouse species. Briefly, mouse sperm heads were isolated from the tail and injected into the mature oocytes. Following injection, the oocytes were thoroughly washed and cultured until day 5 (120 hours) in benchtop incubators at 37°C, 5% CO2, and 5% O2. Embryo development was monitored until the last day of culture, at which point images of each embryo were acquired.

### Clinical Treatment using Fertilo and Traditional IVM

Once informed consent was obtained, subjects were screened and evaluated for participation. If required, patients were pre-treated for vitamin D deficiency or insulin resistance using calcifediol or metformin respectively before beginning stimulation. Eligible subjects that met all of the inclusion criteria were started on oral contraceptive pills (OCPs) for 10-15 days with a 5-day washout period. For evaluative purposes, the final day of the washout period was considered Day 1 of the menstrual cycle. On Day 2, transvaginal ultrasound (TVU) was performed to assess the antral follicle count (AFC) and follicle diameter for monitoring; in addition, baseline evaluation of serum levels of E2, P4, LH, and FSH were performed via a local laboratory. On Day 2, controlled ovarian stimulation (COS) began via administration of 100 mg clomiphene citrate (Clomid) once daily for 5 days (Day 2-Day 6). Ultrasound was performed on Day 7 to determine the number of 150 IU rFSH (Gonal-F) injections that would be administered. If the leading follicle was 6 mm or less, three once daily injections of rFSH were administered on Days 7, 8, and 9. If follicles were 7 mm to 9 mm on Day 7, 150 IU (Gonal-F) was administered once daily on Day 7 and 8, for a total of two injections. If the leading follicle was greater than 9 mm on Day 7, a single injection of 150 IU (Gonal-F) was administered on Day 7.

The ideal leading follicle diameter range was 10-12 mm. On Day 8, 9 or 10, a recombinant hCG (Ovidrel) trigger of 250 µg was administered and the oocyte retrieval (OR) was scheduled for 34-36 hours later. In the event of rapid follicular growth, if a follicle was 13 mm or greater at the Day 2 or Day 7 ultrasound, the cycle was cancelled and repeated in a subsequent menstrual cycle.

In the observational safety phase, all twenty subjects were assigned to OSC-IVM (*Fertilo*) treatment. For the control comparator phase, patients were randomized at a 1:1 ratio to OSC-IVM (*Fertilo*) or Traditional IVM (Media Only IVM) on the day of hCG trigger. Oocyte retrieval was performed according to standard clinical practice using a 17-gauge single lumen needle with 70 mm Hg suction pressure. Rapid needle movement (curettage) was performed to improve oocyte extraction from the follicular wall. All oocyte retrievals were performed under anesthesia. Oocytes were collected into warmed collection media containing heparin in order to minimize clotting. Further, tubing length was minimized and the needle was flushed every 4-5 punctures to minimize clotting and temperature fluctuations during retrieval. No follicular flushing was performed in any retrieval cycle.

Oocyte search was conducted according to standard IVM procedures. Briefly, follicular aspirate was kept warm and passed over a 70 micron cell strainer to separate and filter blood from potential cumulus oocyte complexes (COCs). The filter was then overturned and the COCs were released into a search media lacking heparin (ASP) via dipping and pipette expulsion. COCs were identified under stereomicroscope and transferred to a warmed holding droplet until all oocytes had been found. Typical IVM oocytes were highly compact, containing one to a few layers of cumulus enclosure. Expanded COCs were rarely identified, due to the use of an hCG trigger, and proper monitoring during stimulation was implemented to keep the leading follicle under 12 mm.

All COCs were cultured in either *Fertilo* or Traditional IVM for 30 hours. IVM culture was performed the same for both conditions. Briefly, IVM was performed in Birr 4+8 dishes using microdroplet culture. Culture droplets were 100 µL and were utilized for the culture of up to 5 COCs. Traditional IVM utilized the IVM basal media containing supplementation with 75 mIU/mL rFSH (Gonal-F), 100 mIU/mL hCG (Ovidrel), and 10 mg/mL HSA. For OSC-IVM, the same IVM basal media was utilized and performed as previously described. In OSC-IVM, 100,000 viable OSCs were supplemented into each 100 µL droplet the day of oocyte retrieval. All culture media plates were prepared the day before oocyte retrieval to allow for equilibration under IVF grade oil in 37°C incubators with CO2% to yield a pH of 7.2 to 7.4 and 5% O2. For OSC-IVM, the OSCs (*Fertilo*) were delivered to the clinic under liquid nitrogen vapor shipment and stored in liquid nitrogen until use. For each application, at least two vials of OSCs were prepared to enable treatment of up to 30 COCs across 6 culture droplets, with each droplet containing 100,000 OSCs. Briefly, the day of oocyte retrieval OSCs were thawed at 37°C for 2-3 minutes, gently resuspended using IVM basal media, and centrifuged at 300 x G for 5 minutes to pellet the cells. Supernatent was fully removed to eliminate residual cryoprotectant (Cryostore CS10) and the cells were resuspended using a defined volume of equilibrated complete IVM media described above. A cell count was performed using a cell counting chamber and 100,000 viable OSCs were supplemented into each droplet by replacement of the corresponding volume with the cell suspension. The dishes were incubated for at least 30 minutes prior to oocyte co-culture to allow for re-equilibration. All COCs were washed in IVM basal media and moved into the culture droplets of either OSC-IVM or Traditional IVM and cultured for 30 hours in a 37°C incubator at a %CO2 to achieve a pH of 7.2 to 7.4 and 5% O2.

Mature oocytes were fertilized using ICSI and cultured in group or single culture using single-step embryo culture medium. Assessment of the fertilization rate was performed at 16-18 hours post-ICSI, then at Days 3, 5, 6, and 7 post IVM. A standardized embryo grading system based on the classification system by Gardner & Schoolcraft (Gardner classification) was used on Days 5-7. Embryos that did not meet their respective checkpoints were discarded and blastocysts of freezable quality (Gardner grade 3BB or greater) underwent vitrification (Kitazato). Embryos were hatched using laser-assisted hatching and underwent trophectoderm biopsy prior to vitrification on the day of freezing. Preimplantation genetic testing for aneuploidy (PGT-A) was performed using microarray-based or SNP-based NGS analysis in local laboratories. Only embryos considered euploid were utilized for transfer. Low grade mosaicism, after consultation with a geneticist, were utilized for transfer depending on patient preference. The highest quality embryo was thawed later for frozen embryo transfer (FET) within 2 months of cryopreservation. Only single blastocyst transfers were allowed and all transfers were FET cycles.

For uterine priming, starting on the first day of menses, subjects received 2 mg oral estradiol treatment twice daily with an optional 100 µg patch replaced every three days until the endometrial thickness was ≥7 mm. Progesterone was then administered at 50 mg intramuscular once daily to prepare for ET or via a combination of intravaginal (800 mg) and subcutaneous progesterone. Progesterone could be increased to 100 mg daily if clinically indicated due to bleeding or a low progesterone level.

Following ET, serum beta human chorionic gonadotropin (BHCG) levels were measured 10-14 days post-transfer and followed until an appropriate rise was confirmed (2-3 days following 1st confirmation), indicating a biochemical pregnancy.

Transvaginal ultrasounds were performed at 4-6 weeks gestation to establish the presence of a gestational sac, indicating clinical pregnancy, and a follow-up ultrasound was performed at 8-12 weeks of gestation to confirm ongoing pregnancy with a normal, healthy heartbeat. If a subject became pregnant, P4 and E2 administration were continued for up to 8-12 weeks of gestation.

The site staff followed-up with subjects who had an ongoing pregnancy after discharge from the clinic to obtain a second trimester pregnancy ultrasound record (Week 20-24 Anomaly scan performed by an OBGYN) and serious adverse events (SAE) collection via a second trimester in person visit. Additionally, the clinic obtained birth data from the patient. Adverse events of special interest (AESI) such as response to stimulation and retrieval, ovarian hyperstimulation syndrome (OHSS), pregnancy loss, fetal anomalies, pregnancy complications, birth weight, birth defects, and neonatal complications were collected. For the purposes of this evaluation, only the outcomes of the first embryo transfer were considered in the rates.

### Data Analysis

Significant differences in oocyte maturation rates were evaluated using unpaired t-tests compared to control with a p-value <0.05 considered statistically significant in all cases. For the MOMA, a Fisher’s exact test was used to compare maturation, ICSI survival, cleavage, and blastocyst formation rates. A p-value <0.05 was considered statistically significant in all cases. Data analyses were conducted using GraphPad Prism software (GraphPad Software, San Diego, CA, USA). In the clinical evaluation second phase (comparative), for all demographic comparisons between arms, an unpaired, two-tailed T-test was performed. The comparative statistical analysis between Traditional IVM and *Fertilo* groups for embryological outcomes was performed using logistic regression. Analysis was performed by logistic regression on outcomes per COC per cycle comparing *Fertilo* versus Traditional IVM treatment. Significance was based on the *Fertilo* distribution compared to the Traditional IVM mean for each development point. A p-value of less than 0.05 was considered statistically significant in all tests. Both phases of the evaluation were designed as a small cohort, observational analysis to provide preliminary evidence of safety and efficacy at the embryological level and was not powered for determination of superiority or non-inferiority for *in vivo* endpoints after embryo transfer.

### Data Availability

All data will be made available upon request. For clinical data, appropriate anonymized data will be made upon valid request.

## Supplementary Tables

**S1-** Manifest of Ovarian Support Cell Batch Production and Specifications

**S2-** Factors tested in Design of Experiments Strategy

**S3-** Differentially Expressed Genes per Cluster

**S4-** Proteomics of Ovarian Support Cells (OSC) versus human induced pluripotent stem cells (hiPSC)

**S5-** Secretome of RUO-OSC and CG-OSC

**S6-** Proteomics of Ovarian Support Cells (OSCs) 24 hours in culture versus OSCs 0 hours in culture

**S7-** OSC-IVM lot-based experimental conditions

**S8-** Clinical Patient Demographics 5A

**S9-** Clinical Patient Demographics 5B

