## Supplementary Figures for "Translation of a human induced pluripotent stem cell-derived ovarian support cell product to a Phase 3 enabling clinical grade product for *in vitro* fertilization treatment"

fig. S1: Expression of FOXL2 across different substrates.

fig. S2: Characterization of the candidate clonal CG-hiPSC lines as starting material for OSC differentiation

fig. S3: Residual hiPSC are not detected in the differentiated population of OSCs.

fig. S4: Characterization of specific granulosa cell populations in RUO-OSC-M.

fig. S5: Distribution of granulosa cell markers within the different ovarian support cell (OSC) populations.

fig. S6: MII maturation rates following OSC-IVM with different lots of OSCs.

fig. S7: Expression levels of key growth factors and ligand-receptors in granulosa cells.

fig. S8: Proteomic analysis of RUO- and CG-OSCs and hiPSCs.

fig. S9: Master Cell Bank (MCB) and final Fertilo product manufacturing workflow.

fig. S10: Schematic of the end-to-end chain of custody of the Fertilo product.

fig. S11: Clinical Evaluation Design.

fig. S12: Collected data on adverse events.

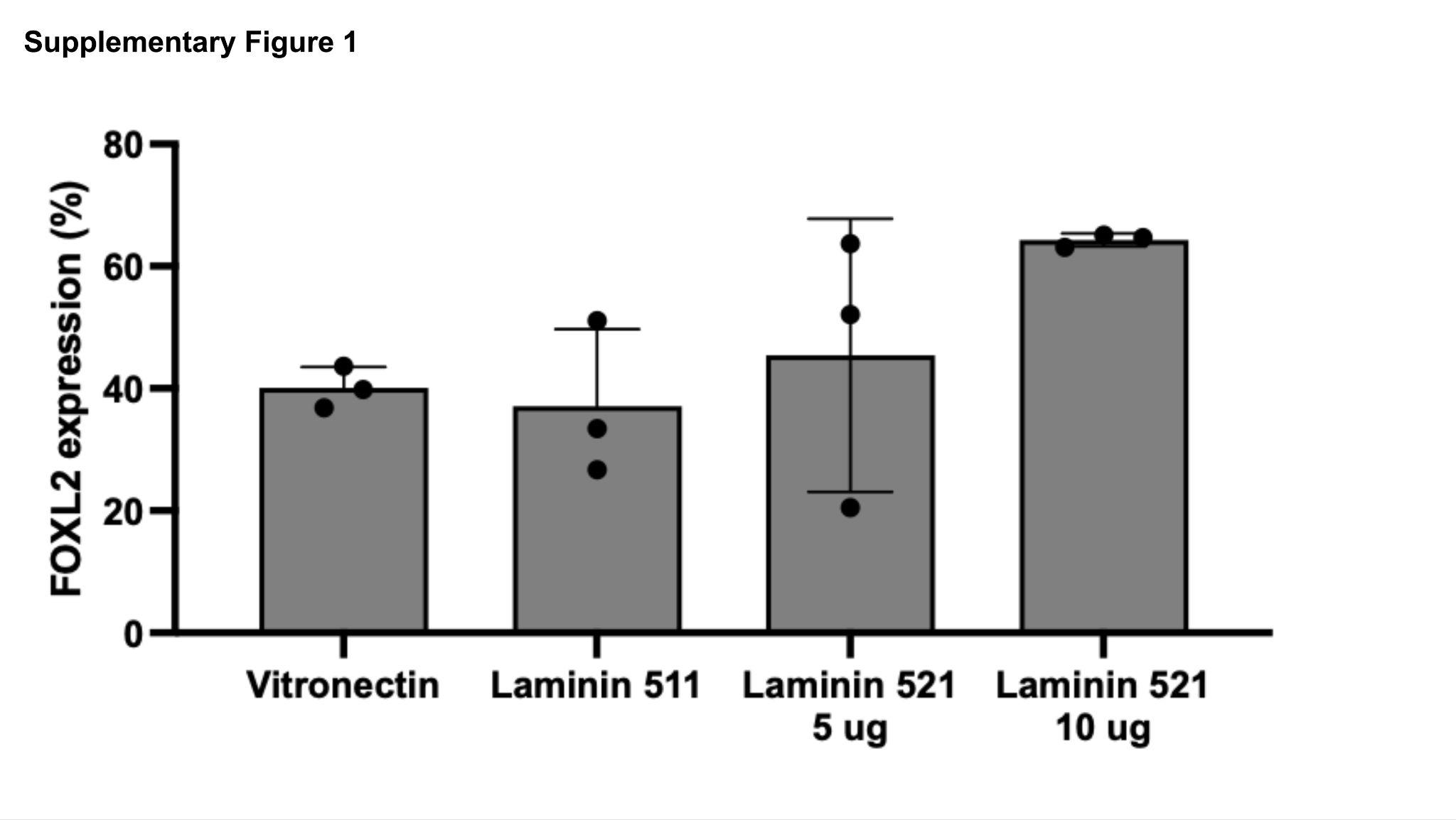

**fig. S1: Expression of FOXL2 across different substrates.** Bar plot demonstrating percentage of FOXL2 expression measured by flow cytometry on day 5 of OSC differentiation onto multiple substrates. Referent to Figure 2.

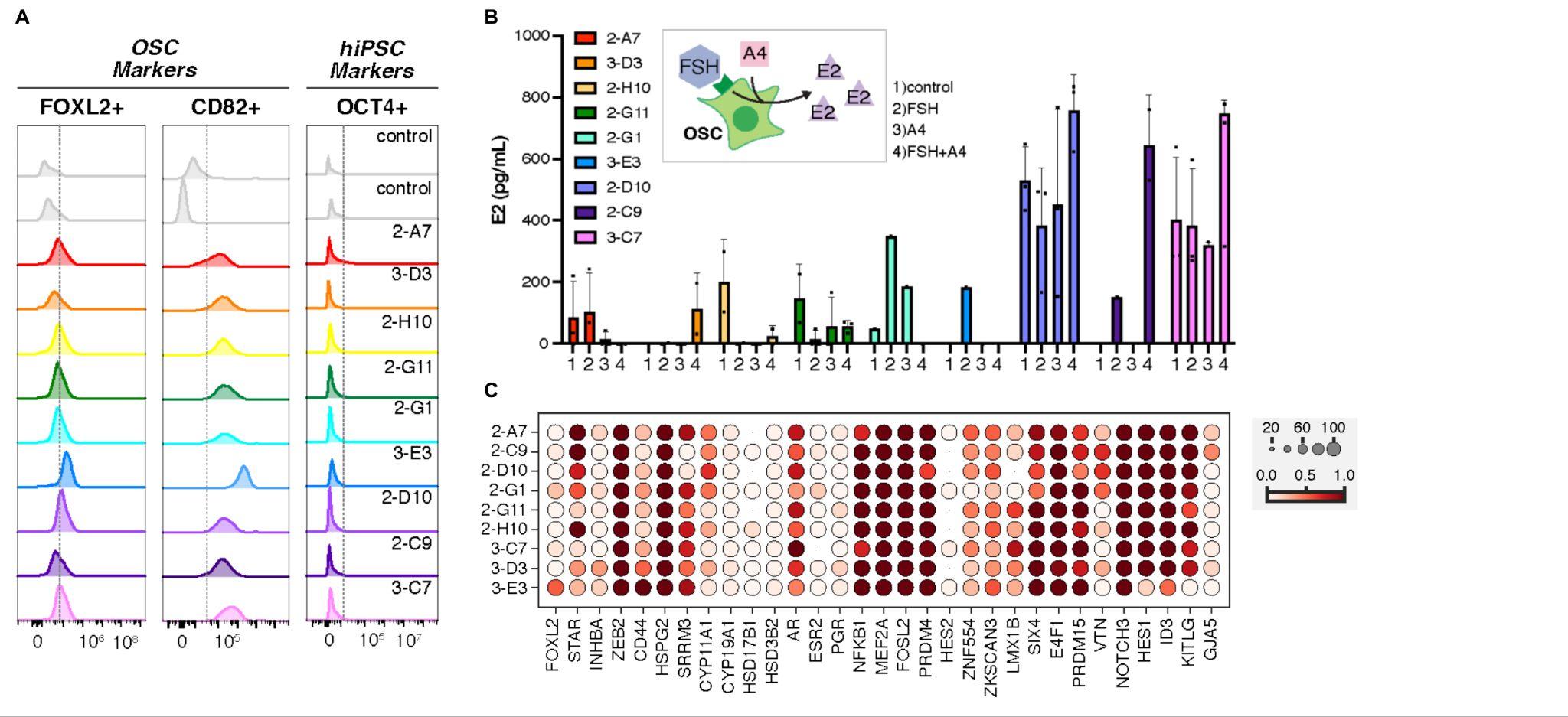

**fig. S2: Characterization of the candidate clonal CG-hiPSC lines as starting material for OSC differentiation.** A) Flow cytometry analysis of two OSC markers, FOXL2+ and CD82+, and the hiPSC marker OCT4+. The expression levels of two controls along with the 9 VCT clones. B) Quantification of estradiol (E2) levels (pg/mL) in conditioned media generated by each clone when cultured in control media (1), control+FSH (2), control+A4 (3), and control+FSH+A4 (4), for 48 hours. Data is the mean ± SEM. C) Dotplot representing the expression of granulosa cell markers in the VCT-clones. Scale represents ‘Mean expression in groups’ ranging from 0 to 1, and circles represent ‘Fraction of cells in group (%) ranging from 0 to 100. Referent to Figure 2.

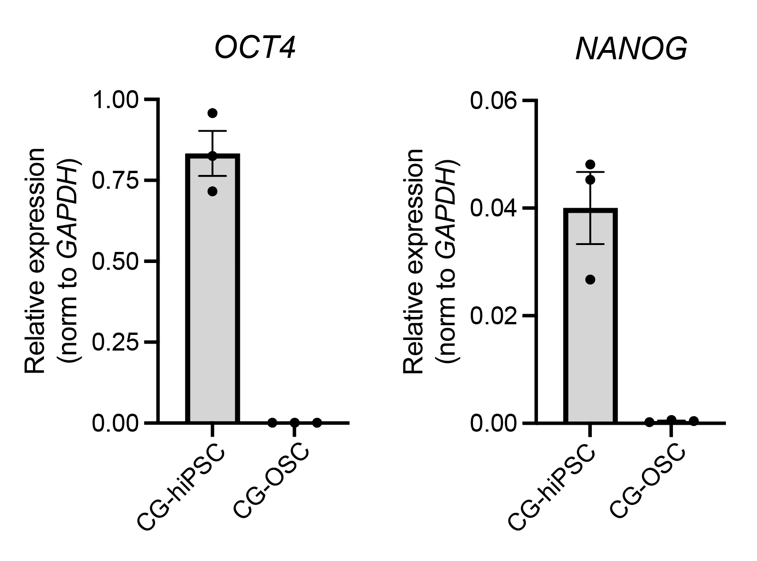

**fig. S3: Residual hiPSC are not detected in the differentiated population of OSCs.** Bar plots depicting relative expression of hiPSC related markers, OCT4 and NANOG in OSCs after 5 days of differentiation (CG-OSCs). Clinical-grade (CG)-hiPSC are used as a positive control. Expression is normalized by expression of the housekeeping gene, GAPDH. Referent to Figure 2.

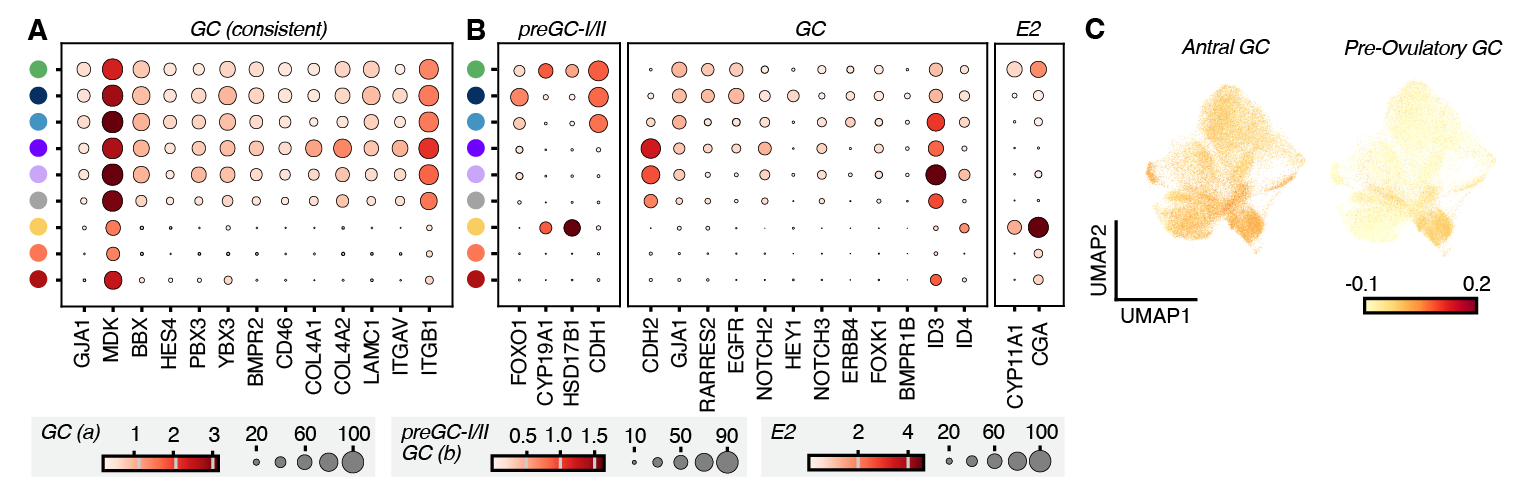

**fig. S4: Characterization of specific granulosa cell populations in RUO-OSC-M.** A) Dotplot representing the expression of granulosa cell marker genes across GC clusters in the RUO-OSC-M subset. Scale represents ‘Mean expression in groups’, ranging from 0 to 3, 0 to 1.5 and 0 to 4, respectively. The circles represent the fraction of cells in the group (%) ranging from 0 to 100, 0 to 90 and 0 to 100, respectively. B) Dotplot representing the expression of preGC-I/II marker genes, granulosa cell marker genes, and steroidogenesis (E2) related genes that supported assignment of each cluster in the RUO-OSC-M subset. Scale represents ‘Mean expression in groups’, ranging from 0 to 3, 0 to 1.5 and 0 to 4, respectively. The circles represent the fraction of cells in the group (%) ranging from 0 to 100, 0 to 90 and 0 to 100, respectively. C) UMAP depicting the signature scores for Antral GC genes and Pre-Ovulatory GC genes. The color scale ranges from -0.1 to 0.2. Referent to Figure 1.

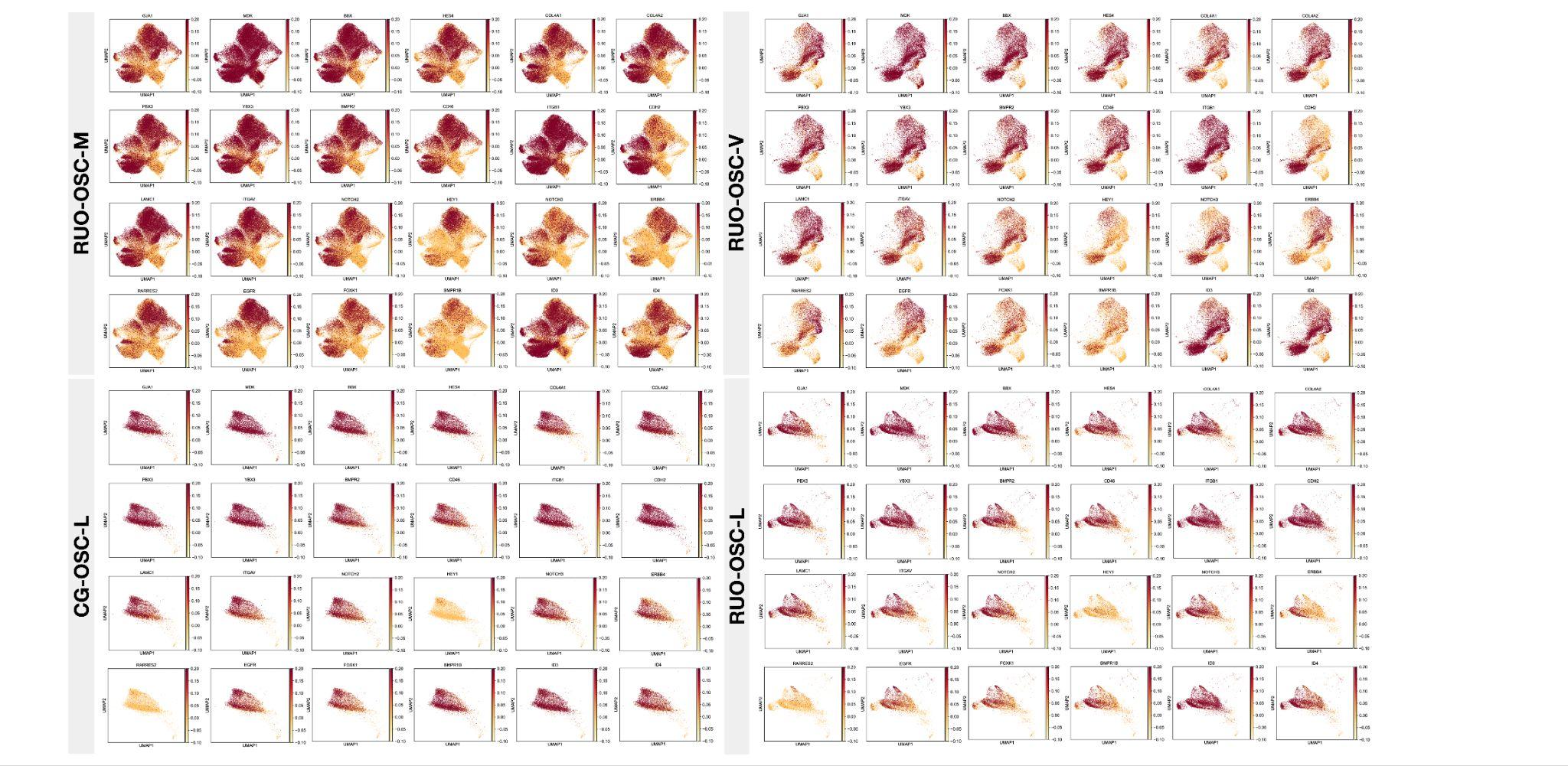

**fig. S5: Distribution of granulosa cell markers within the different ovarian support cell (OSC) populations.** Feature plots depicting expression of granulosa cell marker genes across the RUO-OSC-M subsets, the RUO-OSC-V subsets, the RUO-OSC-L subsets, and the CG-OSC-L subsets. M: Matrigel, V: Vitronectin, L: Laminin. Referent to Figure 3.

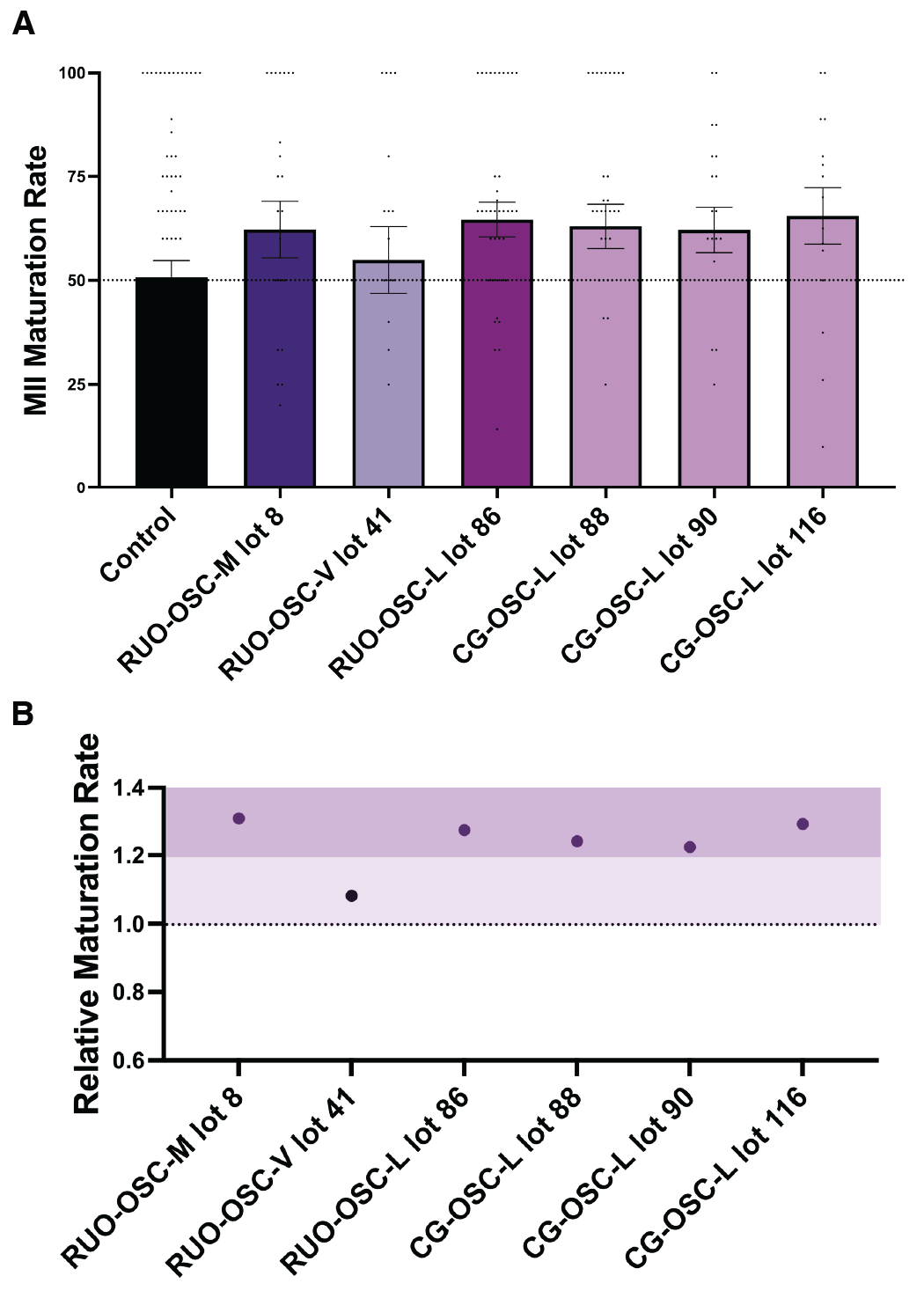

**fig. S6: MII maturation rates following OSC-IVM with different lots of OSCs.**

A) Comparison of MII Maturation Rates between Control-IVM group (grey) and OSC-IVM groups with OSCs from different conditions (RUO-OSC-M, RUO-OSC-V, RUO-OSC-L, and CG-OSC-L). Data is the mean ± SEM (*p*=0.029, RUO-OSC-M lot 8/control: 1.31; *p*=0.018, RUO-OSC-V lot 41/control: 1.08; *p*=0.018, RUO-OSC-L lot 86/control: 1.27; *p*=0.019, CG-OSC-L lot 88/control: 1.24, lot 90/control: 1.22, lot 116/control: 1.29). B) Plot depicting the relative maturation rate per each lot, scale ranging from 0.6 to 1.4. Referent to Figure 3.

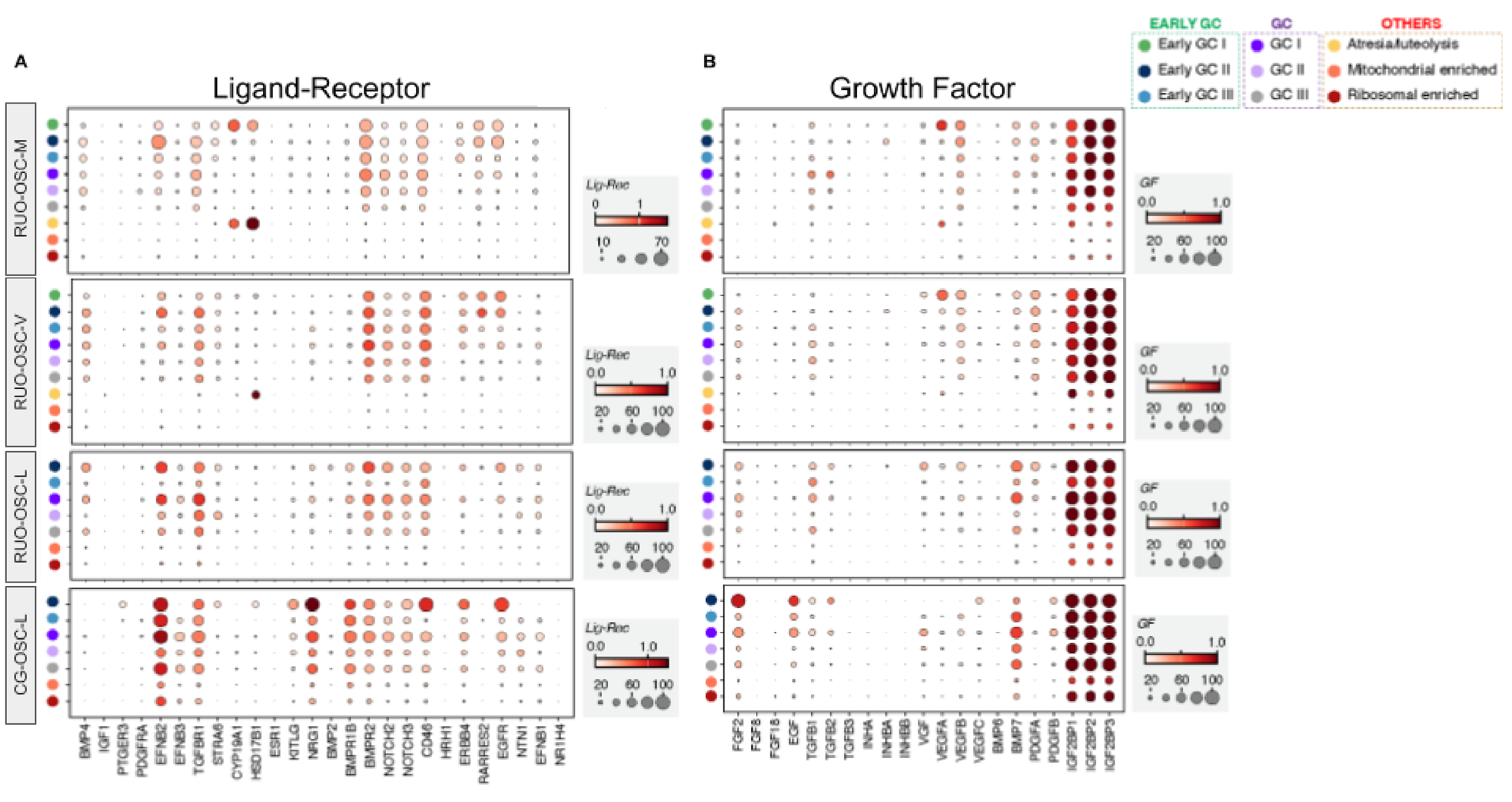

**fig. S7: Expression levels of key growth factors and ligand-receptors in granulosa cells.** A) Dotplot representing the expression of ligand-receptor genes in the RUO-OSC-M, RUO-OSC-V, RUO-OSC-L, and CG-OSC-L subsets. Scale represents ‘mean expression in groups’ ranging from 0 to 1.5, and circles represent ‘fraction of cells in group (%) ranging from 0 to 100. B) Dotplot representing the expression of growth factor genes in the RUO-OSC-M, RUO-OSC-V, RUO-OSC-L, and CG-OSC-L subsets. Scale represents ‘mean expression in groups’ ranging from 0 to 1, and circles represent ‘fraction of cells in group (%) ranging from 0 to 100. Referent to Figure 3.

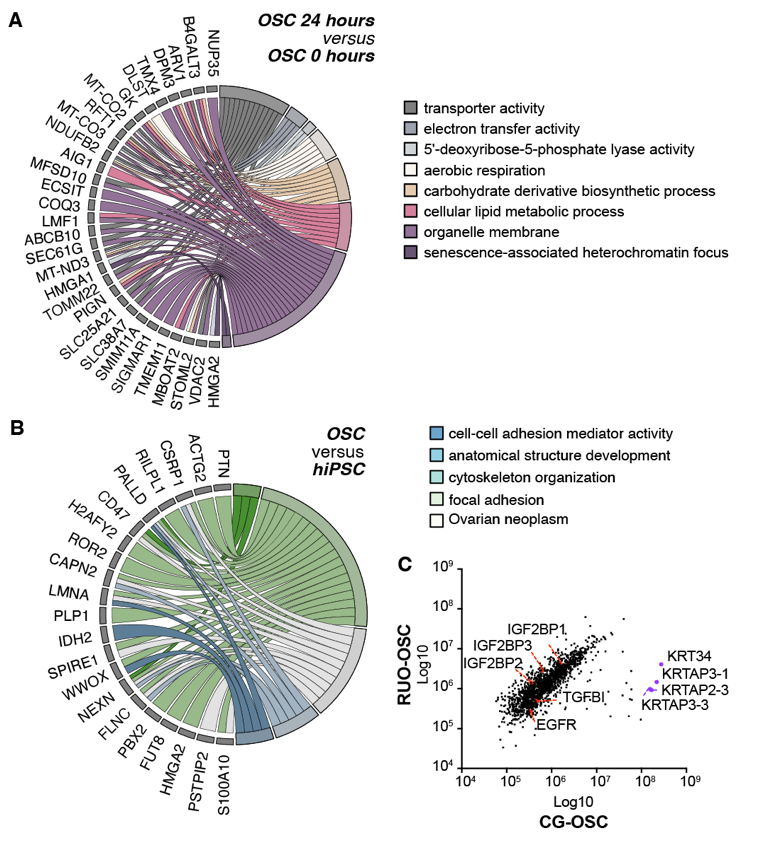

**fig. S8: Proteomic analysis of RUO- and CG-OSCs and hiPSCs.** A) GO chord plots for differently regulated proteins in RUO-OSCs and CG-OSCs at 24 hours versus OSCs at 0 hours. B) GO chord plot for differently regulated proteins in RUO-OSCs and CG-OSCs versus hiPSC. C) Correlation curve for proteins in the secretome of RUO-OSCs versus CG-OSCs. Referent to Figure 3.

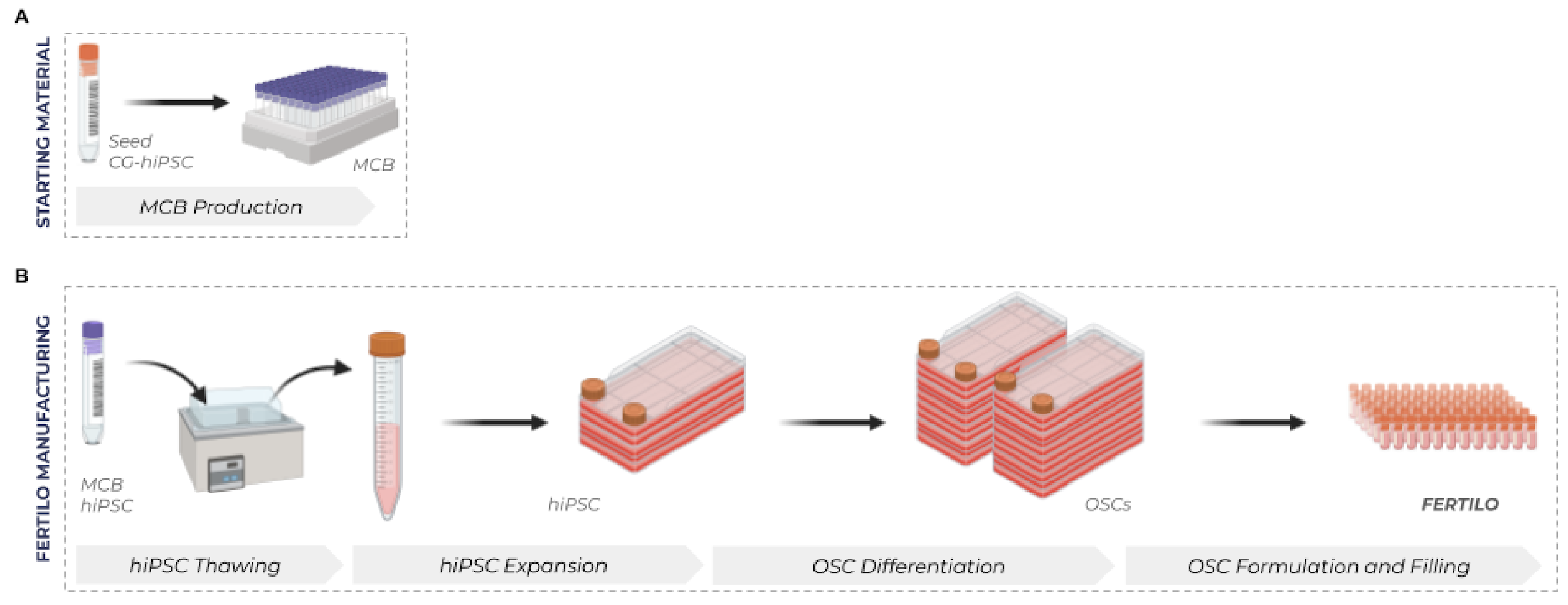

**fig. S9: Master Cell Bank (MCB) and final Fertilo product manufacturing workflow.** A) The CG-hiPSC Seed Bank was expanded into an hiPSC MCB under GMP compliant conditions. B) Workflow for full scale GMP production of the final OSC product (Fertilo).

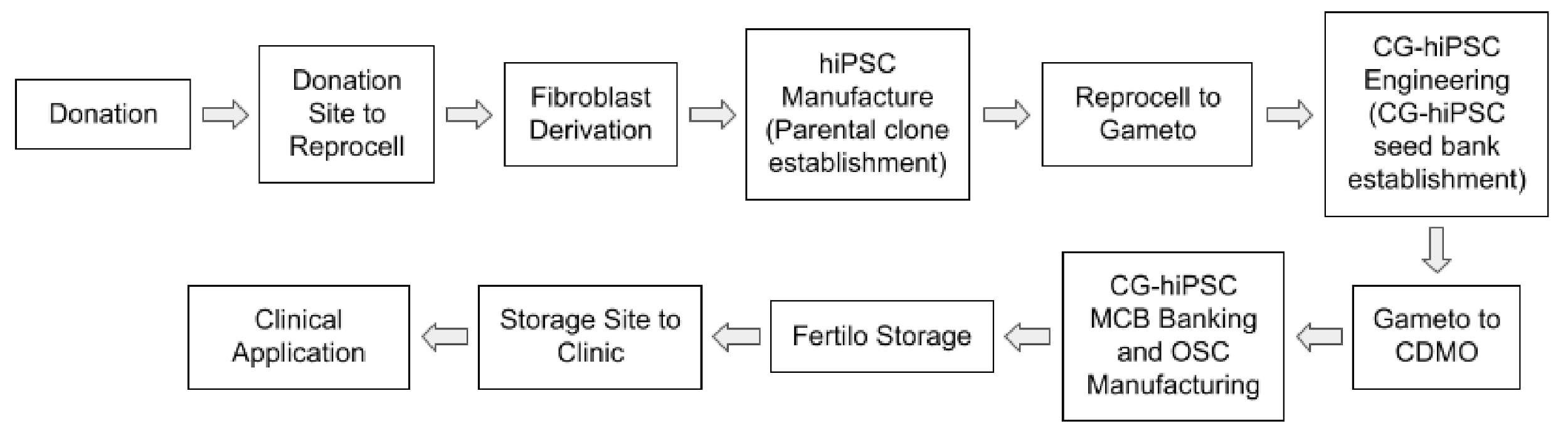

**fig. S10: Schematic of the end-to-end chain of custody of the Fertilo product.** The workflow shows the traceability of the final product from initial parent sample collection through Fertilo product distribution for clinical application.

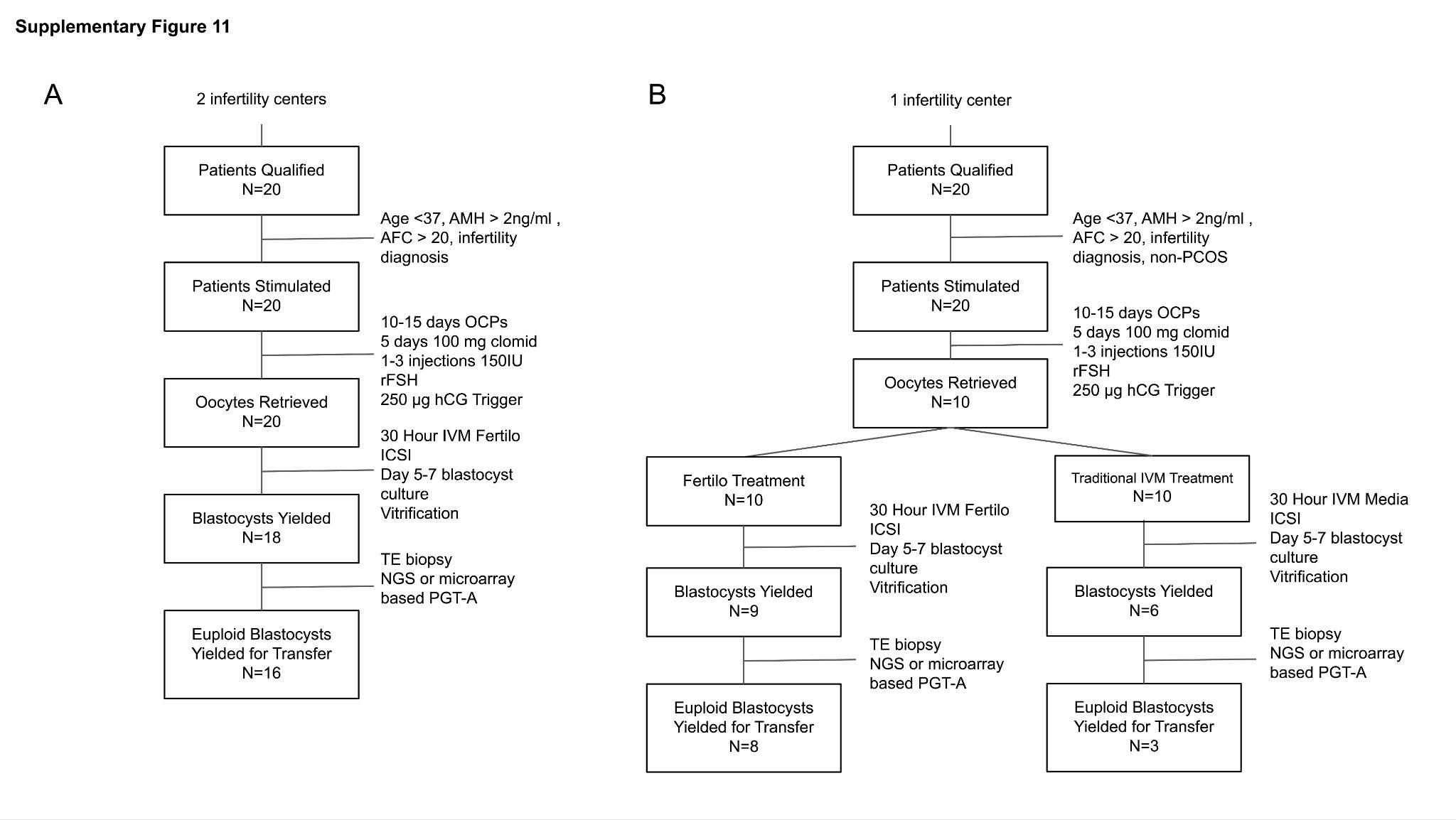

**fig. S11: Clinical Evaluation Design.** A) Workflow of Phase 1 of the clinical evaluation. This phase was a multicenter, single arm observational evaluation in twenty patients with the primary purpose of safety evaluation and establishment of success metrics. B) Workflow of Phase 2 of the clinical evaluation. This phase was a limited comparator controlled cohort evaluation of OSC-IVM (Fertilo) versus traditional IVM (Media Only-IVM) in a single center with the primary purpose of providing measures of efficacy and informing future registrational clinical trial designs. Referent to Figure 5.

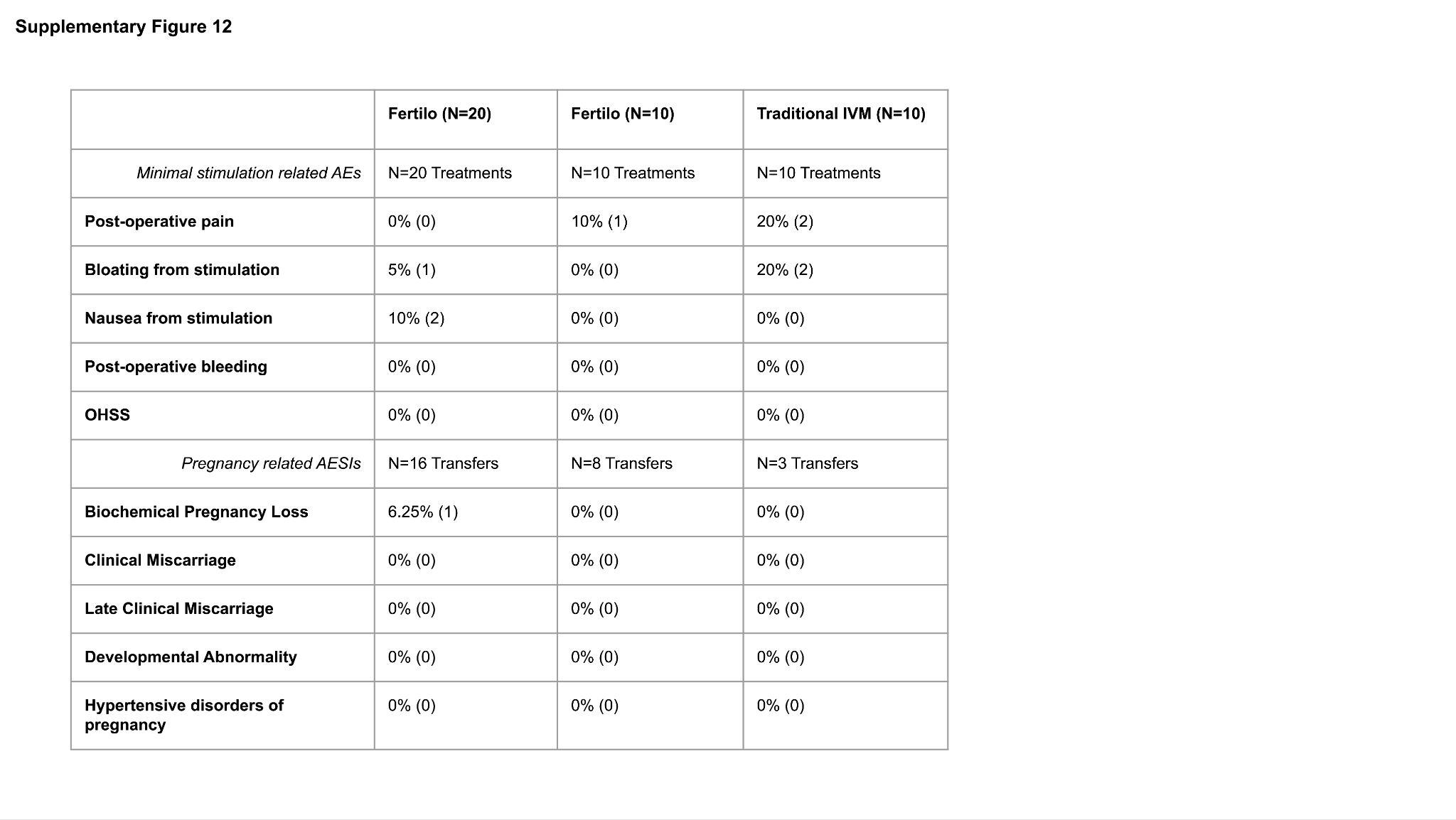

**fig. S12: Collected data on adverse events.** Compiled minimal stimulation-related adverse events (AEs) and pregnancy-related adverse events of special interest (AESIs) for both phases of the clinical evaluation subset by group. The second column corresponds to Phase 1 of the clinical evaluation, and the third and fourth columns correspond to Phase 2 of the clinical evaluation.
